# Moderate to Vigorous Intensity Locomotor Training After Stroke: A Systematic Review and Meta-Analysis of Mean Effects and Individual Response Variability

**DOI:** 10.1101/2022.11.01.22281808

**Authors:** Pierce Boyne, Allison Miller, Owen Kubalak, Caroline Mink, Darcy S. Reisman, George Fulk

## Abstract

**Background and Purpose:** This meta-analysis quantified mean effects of moderate-to-vigorous intensity locomotor training (LT_mv_) on walking outcomes in subacute and chronic stroke, and the magnitude of individual variability in LT_mv_ response.

**Methods:** Databases were searched for randomized trials comparing LT_mv_ with no intervention, non-gait intervention or low-intensity gait training. Comfortable gait speed (CGS), fastest gait speed (FGS), 6-minute walk distance (6MWT), walking activity (steps/day) and adverse effect/event (AE) data were extracted. Pooled estimates were calculated for mean changes within and between groups, the relative risk of different AEs, and the standard deviation of individual response (SD_IR_) to LT_mv_ versus control groups, stratified by study chronicity where possible.

**Results:** There were 19 eligible studies (total N=1,096); 14 in chronic stroke (N=839) and 5 in subacute stroke (N=257). Compared with control interventions, LT_mv_ yielded significantly greater increases in CGS, FGS and 6MWT in both subacute and chronic stroke, with subacute studies showing significantly greater effect sizes for CGS, FGS and nearly 6MWT (p=0.054). In 4 studies reporting steps/day data, LT_mv_ was not significantly different from control interventions. In 14 studies reporting on AEs, there were no treatment-related serious AEs among 398 LT_mv_ participants. SD_IR_ estimates indicated significant individual response variability for CGS, nearly FGS (p=0.0501) and 6MWT.

**Discussion and Conclusions:** LT_mv_ improves mean walking capacity outcomes in subacute and chronic stroke and does not appear to have high risk of serious harm, but response magnitude varies between chronicity subgroups and individuals, and few studies have tested effects on daily walking activity or non-serious AEs.

## 1. INTRODUCTION

Clinical practice guidelines recommend moderate-to-vigorous intensity locomotor training (LT_mv_) to improve walking speed and distance among persons with chronic stroke (>6 months post).^1^ However, the effects of this approach have not been previously synthesized in subacute stroke, nor have they been quantitatively pooled with meta-analysis. Modern meta-analysis can improve the precision of effect estimation (narrower confidence intervals) by combining results across studies, and can support more valid inference than other synthesis methods.^2^ Meta-analysis can also estimate the *magnitude* of treatment-related improvement on the scale of the original measurement, which facilitates interpretation of clinical meaning^2^ and could inform LT_mv_ goal setting and progress monitoring.

While meta-analysis is most commonly used to pool mean treatment effects, it can also be used to pool estimates of response variability.^3^ This could be helpful to determine the need for prognostic studies of treatment response, since individual response prediction is only relevant if there is substantial response variability.^4^ Prior LT_mv_ studies have already begun exploring predictors of treatment response.^e.g. 5-12^ However, no previous studies have assessed the true amount of individual variability in LT_mv_ response, while controlling for variability due to measurement error.

Intervention studies in rehabilitation and other fields often appropriately report measures of within-group change variability (e.g. standard deviation of change within the experimental group).^4,13,14^ But these cannot be interpreted as measures of response variability because they will always be inflated by measurement error, to a varying degree depending on the test-reliability of the outcome measure in the study sample.^4,13,14^ Thus, within-group estimates of change variability alone will always overestimate the true variability in treatment response.

The standard deviation of individual response (SD_IR_) overcomes this limitation by calculating response variability in the experimental group relative to a control group that is also subject to the same measurement error.^4,13,14^ Thus, an SD_IR_ greater than zero indicates true variability in response to an intervention, and the magnitude of the SD_IR_ estimates how much individual responses truly vary from the mean response. This could inform clinicians and patients/clients about the likely range of possible outcomes to expect from an intervention and the probability of a meaningful improvement.^4,13,14^ However, no previous studies have estimated the SD_IR_ for LT_mv_. Also, few previous meta-analyses in any field have synthesized SD_IR_ estimates across studies,^15,16^ and this is a promising strategy to overcome the lower precision and statistical power when estimating measures of variability (e.g. SD_IR_) rather than means.^3^

The current meta-analysis aimed to quantify mean effects of LT_mv_ on walking outcomes in subacute and chronic stroke, the risk of harms and the magnitude of true individual variability in LT_mv_ response. Using meta-regression,^17,18^ we also tested associations between effect sizes and study characteristics, including participant features, treatment parameters and risk of bias.

## 2. METHODS

### 2.1. Search Strategy

Studies were identified by searching electronic databases and scanning reference lists of articles from January to April of 2022. The database search included Pubmed, MEDLINE, Cumulative Index of Nursing and Allied Health Literature (CINAHL), Physiotherapy Evidence Database (PEDro), and Academic Search Complete. Keywords and combinations are shown in the Appendix. This review was not pre-registered.

### 2.2. Eligibility Criteria

#### Types of studies

Randomized studies comparing LT_mv_ to a control group involving no intervention, non-gait intervention or low-intensity gait training, among stroke survivors. Reports had to be published in a peer-reviewed journal from 1/1/1995 to 3/1/2022 and the full text had to be available in English. When the same study was reported in multiple publications, the most complete report for each outcome was used for extraction.

#### Types of participants

Adult humans with a history of stroke. No restrictions were imposed based on stroke characteristics.

#### Types of interventions

LT_mv_ was defined as repetitive walking practice on a treadmill or overground with no more assistance than needed to enable safe practice (not to optimize kinematics), with either:

1) A moderate or higher mean aerobic intensity (>39% of heart rate reserve [HRR], peak oxygen consumption rate or peak work rate, or >63% of maximal heart rate);^19,20^ or

2) A focus on active training at faster than comfortable speed.

To be eligible for analysis, an LT_mv_ protocol had to include multiple training sessions, all intervention components had to be within the physical therapy scope of practice and an LT_mv_ group could not receive any additional primary intervention that the comparison group did not also receive (e.g. neuromuscular electrical stimulation, brain or spinal cord stimulation, rhythmic auditory stimulation, virtual reality, dual-task training, split-belt dual-speed training, resistance training, circuit training). However, the following did not count as additional primary interventions:

1. Assistance required to enable safe walking practice (e.g. fall protection harness, minimum necessary body weight support, minimum necessary physical or mechanical assist);
2. Additional challenges to augment the intensity or difficulty of walking practice (e.g. incline/stair walking, resisted gait, limb weights, obstacles, gait perturbations, multi-directional gait, dual-task practice that only appeared to take up a small proportion of the experimental treatment time); or
3. Non-gait task practice that only appeared to take up a small proportion of the experimental treatment time.

#### Types of outcome measures

Walking capacity measures included comfortable gait speed (CGS) or fastest gait speed (FGS) over a short distance (e.g. 10m walk test), or a timed walking distance test (e.g. 6-minute walk test [6MWT]). Another outcome of interest was daily walking activity (mean steps per day), measured by an activity monitor. Only change scores from baseline to immediately post-intervention were included, because longer-term follow up time points are less consistent in the literature and more prone to missing data.

### 2.3. Eligibility Determination, Risk of Bias Assessment and Data Extraction

Two authors (OK and CM) independently assessed study eligibility, one author (AM) assessed each included outcome comparison with the Cochrane risk of bias 2 (RoB-2) instrument^2,21^ and another author (PB) performed RoB-2 assessment for adverse effect/event measures and resolved discrepancies and uncertainties. Data extraction was primarily performed by AM and PB, including: 1) study design and eligibility criteria; 2) characteristics of trial participants, like mean stroke chronicity (classified as subacute [< 6 months] or chronic [≥ 6 months]), ambulatory status (whether or not participants could walk short distances without continuous physical assistance from another person), and mean baseline outcome variables; 3) characteristics of the LT_mv_ and control interventions, like control type (intervention targeting lower limb function or not), LT_mv_ modes (treadmill and/or overground), mean LT_mv_ intensity (classified as moderate or vigorous),^19,20^ total LT_mv_ volume in hours, and whether participants were concurrently receiving usual care physical therapy; 4) outcomes, including mean changes and standard deviations of change in each group, between-group differences in mean change and corresponding variances; 5) adverse events in each group; and 6) overall risk of bias (low/some or high) for each comparison, which was categorized as high if more than one of the RoB-2 domains was rated high risk or if all five domains were rated at least some risk.

When mean LT_mv_ intensity was not reported, it was estimated from data provided, including peak intensity, starting intensity target and final intensity target. Moderate intensity was defined as 40-59% HRR or VO_2peak_ or 64-76% HR_max_.^19,20^ Vigorous intensity was defined as 60-84% HRR or VO_2peak_ or 77-93% HR_max_.^19,20^

#### Outcome data extraction

Outcome data extraction used the methods described by the Cochrane Collaborative for mean differences,^2^ prioritizing extraction of within-group mean changes and corresponding standard deviations of change (SDΔ), standard errors, confidence intervals, p values or T statistics where available (in order of priority). When those were not available, we directly extracted estimates of the between-group difference in mean change with corresponding standard errors, confidence intervals, p values, T statistics or F statistics (in priority order). When multiple LT_mv_ or control groups in the same trial were eligible for analysis, we avoided data duplication by pooling data from those groups together with exact averaging weighted by group sample size.^2^ For crossover studies, we extracted data from the initial parallel phase before any crossover.

When the best available measure to calculate variability was a p value inequality (e.g. p<0.05), the highest possible p value with one additional digit (e.g. p=0.049) was used to approximately impute the highest variance (uncertainty) that would be consistent with the report.^22^ When the only available variability estimate was the standard deviation at baseline and post-intervention in each group, we calculated estimates for SDΔ using the minimum repeated measures correlation from other eligible studies that reported all needed data.^2^ Smaller repeated measures correlations are more conservative because they result in higher estimates for SDΔ, reflecting greater uncertainty. These repeated measures correlations for the LT_mv_ and control groups (respectively) were: CGS, 0.70 and 0.62; FGS, 0.69 and 0.83; 6MWT, 0.65 and 0.88.

#### Adverse event data extraction

An adverse event was defined as an undesirable change in health status in a study participant that may or may not be related to study procedures, whereas an adverse *effect* was defined as an adverse event that had a reasonable possibility of being caused by the study intervention (e.g. occurred during training).^2^ Adverse effects of interest were the number of participants experiencing any serious adverse event related to the study intervention^23^ and the number of participants with new or increased musculoskeletal pain during training. The adverse event of interest was the number of randomized participants who experienced falls during study participation. We assumed that there were zero serious adverse effects in both groups if a study reported that no adverse events occurred. However, we did not make this assumption if adverse events were not mentioned, nor did we assume zero musculoskeletal pain effects or fallers unless those events were specifically reported.^2^ If events were not reported by group they were recorded as missing.

#### Estimation of the standard deviation of individual response (SD_IR_) from individual trials

SD_IR_ was calculated for studies that reported separate estimates of change variability for each group. Since standard deviations like SD_IR_ have poor statistical properties,^16^ analyses were done using SD_IR2_ (the variance of individual response) and its standard error:^3,4,13-16^

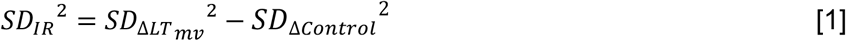

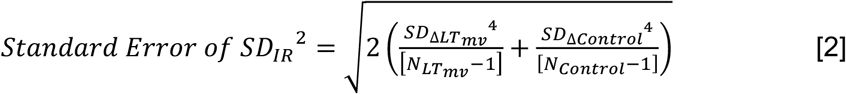

Resulting estimates and confidence intervals were then converted to standard deviation units by taking the square root, while preserving any negative values.^14-16^

### 2.4. Statistical Methods

For outcome data, separate random-effects meta-analyses were performed for each estimate of interest (LT_mv_ – control mean change difference, SD_IR2_ and LT_mv_ within-group mean change) and for each outcome measure, using R^24^ and the ‘meta’ package,^25^ with restricted maximum likelihood estimation of the between-study variance and Hartung-Knapp correction.

For adverse event data, separate random-effects meta-analyses estimated the relative risks (LT_mv_ / control incidence proportions) of each event/effect of interest (treatment-related serious adverse effects, treatment-related musculoskeletal pain and falls), using the ‘meta’ package,^25^ with the exact Mantel-Haenszel method, restricted maximum likelihood estimation of the between-study variance and treatment arm continuity correction in studies with zero cell frequencies when calculating individual study results.

#### Between-study heterogeneity, meta-regression and assessment of publication bias

Between-study heterogeneity was assessed with I^2^, random effects model estimates of the between-study standard deviation (τ) and p-values from the Cochran’s Q test. Generally, I^2^ values less than 30-40% may represent trivial heterogeneity, values between ∼30-60% may represent moderate heterogeneity, and higher values may represent substantial heterogeneity.^2^ Meta-regression models were attempted to explain some between-study variability by testing the association between study effect size and each independent variable of interest, including: stroke chronicity (subacute vs chronic), ambulatory status (all participants ambulatory vs some non-ambulatory), baseline level of the dependent variable, LT_mv_ mode (some overground training vs treadmill only), LT_mv_ intensity (vigorous vs moderate), LT_mv_ volume, control intervention (including vs not including any gait training), and risk of bias (high vs low/some). These meta-regressions were only attempted when there were at least 10 available studies and (for categorical independent variables) at least 3 studies in each category.^2^ When there were sufficient studies, the primary analysis was stratified by stroke chronicity (the first meta-regression variable above) based on the aims of this study and established differences in effect sizes between subacute and chronic stroke studies.^26^ Additional meta-regressions were only attempted within chronicity subgroups to avoid likely confounding.

The risk of publication bias was visually assessed with funnel plots and statistically estimated with Egger’s test (for the continuous outcomes) or the score test (for the dichotomous adverse event data) when there were at least 10 studies.^2^

#### Between-group heterogeneity of (within-study) variance

It is common in meta-analysis to pool outcome variability estimates across groups within the same study, under the assumption that variability (in this case SD_Δ_) does not differ between groups.^2^ Our SD_IR_ analysis directly tested that assumption, since an SD_IR_ significantly different from zero indicates a significant between-group difference in SD. The SD_IR_ results suggested that the homogeneity of variance assumption was not plausible. Thus, we used separate SD_Δ_ estimates for each group in the meta-analyses involving mean changes. For studies where SD Δ estimates were not available by group, we calculated them based on the between-group pooled variance estimate (*SD*_Δpooled_^2^) from that study and the overall SD_IR_ estimate pooled across studies for that outcome, using the formulas:

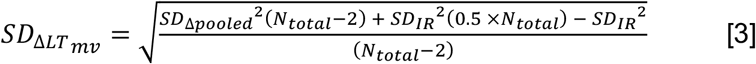

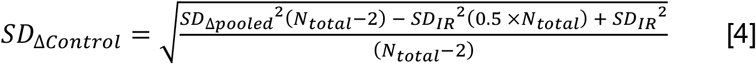

When calculating the standard errors of the between-group differences in mean change, we were then able to use the heterogeneous variance formula [5] rather than the standard homogeneous variance formula [6]:

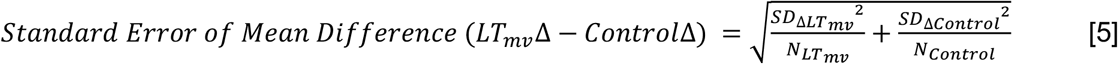

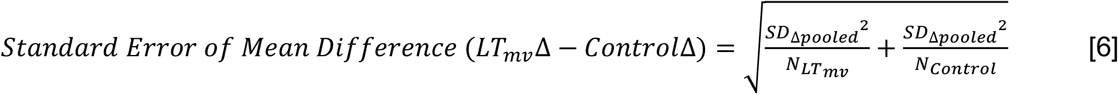

This heterogeneous variance formula [5] is more conservative in that it results in similar or higher standard error (uncertainty) than formula [6], depending on the magnitude of between-group imbalance in SD_Δ_.

#### Imputation of missing outcome data

When a study did not provide results from an intent-to-treat analysis (i.e. when one or more randomized participants were not included in the analyses), missing outcome data were imputed using “strategy 1” from Ebrahim et al.^27^ For participants with missing data, mean change was set at the value in the control group from the same study, and SD_Δ_ was set at the median control group SD_Δ_ across studies for that outcome measure (CGS, 0.09 m/s; FGS, 0.12 m/s; 6MWT, 32.6 m; daily walking activity, 860.5 steps/day). These values were calculated across subacute and chronic stroke studies because median control group SD_Δ_ was similar or identical between chronicity subgroups. The total mean change and SD_Δ_ scores within each treatment group for the full randomized (intent-to-treat) sample were estimated by combining the observed and imputed values using exact weighted averaging.^2^ When calculating standard errors, participants with imputed data were not counted in the sample size to account for uncertainty in the imputed values and avoid artificial narrowing of the confidence intervals. Missing data imputation was not done for SD_IR_ analysis because it would bias the results towards larger SD_IR_, since missing data were imputed with control group means. Missing data imputation was also omitted for adverse event analysis because it would tend to be anti-conservative by shrinking relative risk estimates towards no between-group difference.

#### Estimated proportions of responders

To estimate the proportion of participants with meaningful CGS, FGS, and 6MWT responses attributable to LT_mv_, we modeled the distribution of individual net changes (i.e. individual change with LT_mv_ above and beyond the expected change with control interventions) as a normal curve. Separate distributions were generated for each outcome within each stroke chronicity subgroup. The mean of each distribution was set as the pooled estimate of the mean change difference (LT_mv_ – control) from the primary meta-analysis, and the SD was set as the pooled SD_IR_ (the estimate of the true individual variability in responsiveness).^4,14^ We then calculated the proportion of change differences in this modeled distribution that were above clinically important difference (CID) thresholds reported in the literature. CID thresholds ranged from 0.10 to 0.20 m/s for gait speed^28-31^ and from 14 to 50 m for the 6MWT.^31-33^

We also estimated the proportion of participants with meaningful changes during LT_mv_ (but not necessarily attributable to LT_mv_) by setting the mean of each distribution as the pooled estimate of mean change within LT_mv_ groups, rather than the mean change difference between groups.^4^ This was done to provide estimates that may be more interpretable in the context of clinical practice, where individual patients/clients can be measured before and after LT_mv_, but cannot perform both LT_mv_ and a control intervention in parallel.

## 3. RESULTS

### 3.1. Literature search results

The literature search identified 19 eligible studies (Supplemental Figure S1), with a combined sample size of 1,096 randomized participants (Table 1).^7,8,12,34-49^ Among these studies, 14 were in chronic stroke (N=839) and 5 were in subacute stroke (N=257). For two studies, additional data for outcomes of interest were found in other reports.^50,51^ Our literature search databases are publicly available (title/abstract screening: https://sysrev.com/u/5631/p/93708; full-text review: https://sysrev.com/u/5631/p/103118).

**Table 1.**
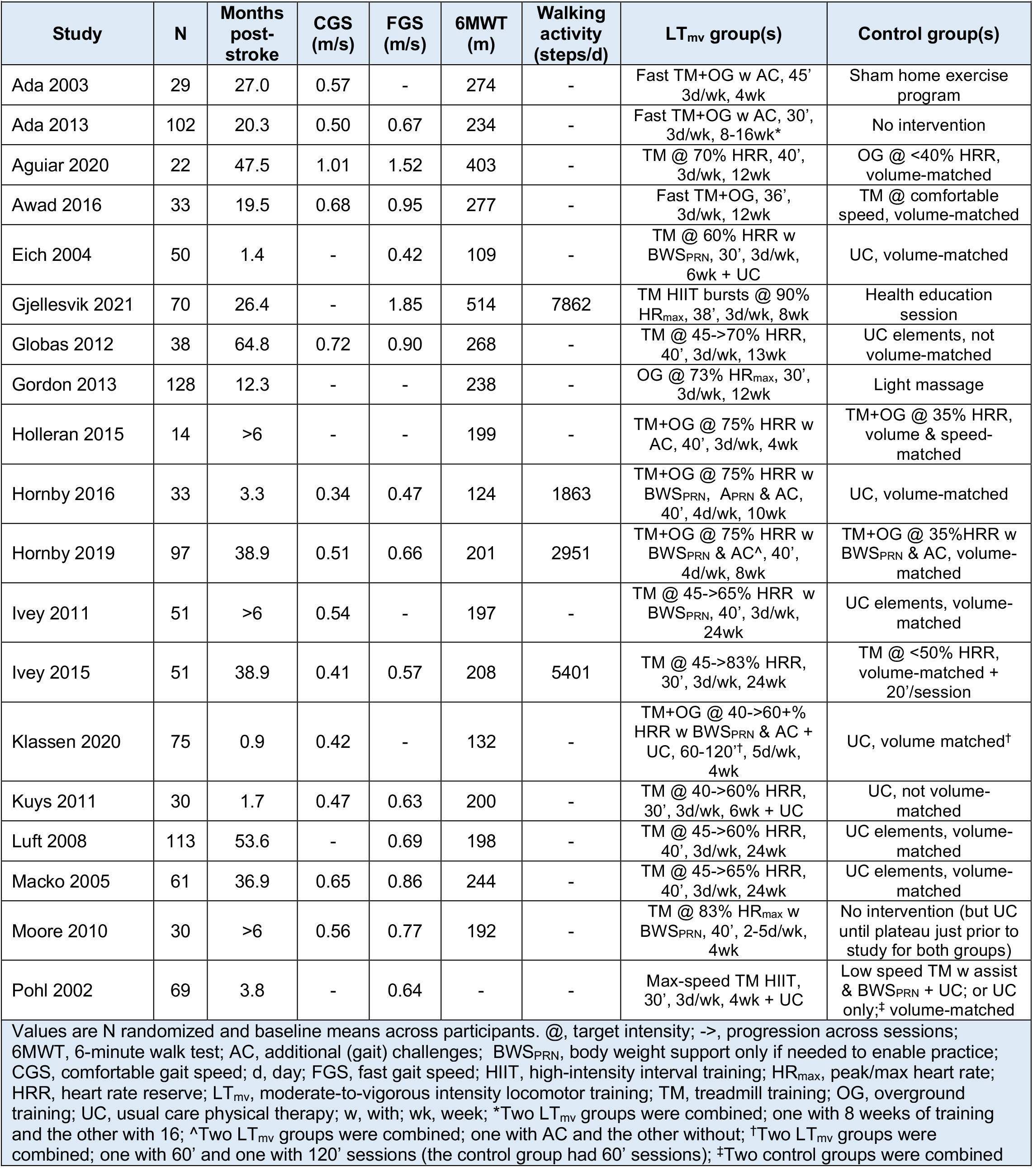
Characteristics of Included Studies.

| Study | N | Months post-stroke | CGS (m/s) | FGS (m/s) | 6MWT (m) | Walking activity (steps/d) | LT<sub>mv</sub> group(s) | Control group(s) |
| --- | --- | --- | --- | --- | --- | --- | --- | --- |
| Ada 2003 | 29 | 27.0 | 0.57 | - | 274 | - | Fast TM+OG w AC, 45' 3d/wk, 4wk | Sham home exercise program |
| Ada 2013 | 102 | 20.3 | 0.50 | 0.67 | 234 | - | Fast TM+OG w AC, 30', 3d/wk, 8-16wk* | No intervention |
| Aguiar 2020 | 22 | 47.5 | 1.01 | 1.52 | 403 | - | TM @ 70% HRR, 40', 3d/wk, 12wk | OG @ <40% HRR, volume-matched |
| Awad 2016 | 33 | 19.5 | 0.68 | 0.95 | 277 | - | Fast TM+OG, 36', 3d/wk, 12wk | TM @ comfortable speed, volume-matched |
| Eich 2004 | 50 | 1.4 | - | 0.42 | 109 | - | TM @ 60% HRR w BWS<sub>PRN</sub>, 30', 3d/wk, 6wk + UC | UC, volume-matched |
| Gjellesvik 2021 | 70 | 26.4 | - | 1.85 | 514 | 7862 | TM HIIT bursts @ 90% HR<sub>max</sub>, 38', 3d/wk, 8wk | Health education session |
| Globas 2012 | 38 | 64.8 | 0.72 | 0.90 | 268 | - | TM @ 45->70% HRR, 40', 3d/wk, 13wk | UC elements, not volume-matched |
| Gordon 2013 | 128 | 12.3 | - | - | 238 | - | OG @ 73% HR<sub>max</sub>, 30', 3d/wk, 12wk | Light massage |
| Holleran 2015 | 14 | >6 | - | - | 199 | - | TM+OG @ 75% HRR w AC, 40', 3d/wk, 4wk | TM+OG @ 35% HRR, volume & speed-matched |
| Hornby 2016 | 33 | 3.3 | 0.34 | 0.47 | 124 | 1863 | TM+OG @ 75% HRR w BWS<sub>PRN</sub>, A<sub>PRN</sub> & AC, 40', 4d/wk, 10wk | UC, volume-matched |
| Hornby 2019 | 97 | 38.9 | 0.51 | 0.66 | 201 | 2951 | TM+OG @ 75% HRR w BWS<sub>PRN</sub> & AC<sup>^</sup>, 40', 4d/wk, 8wk | TM+OG @ 35%HRR w BWS<sub>PRN</sub> & AC, volume-matched |
| Ivey 2011 | 51 | >6 | 0.54 | - | 197 | - | TM @ 45->65% HRR w BWS<sub>PRN</sub>, 40', 3d/wk, 24wk | UC elements, volume-matched |
| Ivey 2015 | 51 | 38.9 | 0.41 | 0.57 | 208 | 5401 | TM @ 45->83% HRR, 30', 3d/wk, 24wk | TM @ <50% HRR, volume-matched + 20'/session |
| Klassen 2020 | 75 | 0.9 | 0.42 | - | 132 | - | TM+OG @ 40->60+% HRR w BWS<sub>PRN</sub> & AC + UC, 60-120'<sup>†</sup>, 5d/wk, 4wk | UC, volume matched<sup>†</sup> |
| Kuys 2011 | 30 | 1.7 | 0.47 | 0.63 | 200 | - | TM @ 40->60% HRR, 30', 3d/wk, 6wk + UC | UC, not volume-matched |
| Luft 2008 | 113 | 53.6 | - | 0.69 | 198 | - | TM @ 45->60% HRR, 40', 3d/wk, 24wk | UC elements, volume-matched |
| Macko 2005 | 61 | 36.9 | 0.65 | 0.86 | 244 | - | TM @ 45->65% HRR, 40', 3d/wk, 24wk | UC elements, volume-matched |
| Moore 2010 | 30 | >6 | 0.56 | 0.77 | 192 | - | TM @ 83% HR<sub>max</sub> w BWS<sub>PRN</sub>, 40', 2-5d/wk, 4wk | No intervention (but UC until plateau just prior to study for both groups) |
| Pohl 2002 | 69 | 3.8 | - | 0.64 | - | - | Max-speed TM HIIT, 30', 3d/wk, 4wk + UC | Low speed TM w assist & BWS<sub>PRN</sub> + UC; or UC only;<sup>‡</sup> volume-matched |

Values are N randomized and baseline means across participants. @, target intensity; ->, progression across sessions; 6MWT, 6-minute walk test; AC, additional (gait) challenges; BWS<sub>PRN</sub>, body weight support only if needed to enable practice; CGS, comfortable gait speed; d, day; FGS, fast gait speed; HIIT, high-intensity interval training; HR<sub>max</sub>, peak/max heart rate; HRR, heart rate reserve; LT<sub>mv</sub>, moderate-to-vigorous intensity locomotor training; TM, treadmill training; OG, overground training; UC, usual care physical therapy; w, with; wk, week; \*Two LT<sub>mv</sub> groups were combined; one with 8 weeks of training and the other with 16; ^Two LT<sub>mv</sub> groups were combined; one with AC and the other without; †Two LT<sub>mv</sub> groups were combined; one with 60' and one with 120' sessions (the control group had 60' sessions); ‡Two control groups were combined

### 3.2. LT_mv_ versus control mean change differences

For the analysis of mean change differences, the three walking capacity outcomes (CGS, FGS and 6MWT) each showed significantly greater improvement with LT_mv_ versus control interventions in both subacute and chronic stroke (Table 2, Fig 1, Figs S2-S3). Subacute studies had significantly greater effect sizes than chronic studies for each of these outcomes except the 6MWT, which still showed a similar tendency (p=0.054). For daily walking activity, there were insufficient studies to stratify the analysis by study chronicity and no significant differences between LT_mv_ and control interventions (Fig S4).

**Table 2.** LT_mv_-Control Mean Change Differences.

|  |  | Mean Change Difference |  | Between-study heterogeneity |  |  |  |
| --- | --- | --- | --- | --- | --- | --- | --- |
| | N studies (total N) | Estimate [95%CI] | P-value | SD ($\tau$) | I<sup>2</sup> | P-value* | R<sup>2</sup> |
| **Comfortable gait speed, m/s** | 13 (652) | 0.08 [0.04, 0.12] | 0.0017 | 0.054 | 67.6% | 0.0002 | - |
| Chronic stroke studies | 10 (514) | 0.06 [0.01, 0.10] | 0.0173 | 0.044 | 65.1% | 0.0022 | - |
| Subacute stroke studies | 3 (138) | 0.16 [0.12, 0.19] | 0.0022 | 0.044 | 0.0% | 0.9144 | - |
| *Subacute vs chronic stroke* | *13 (652)* | *0.10 [0.01, 0.18]* | *0.0263* | *0.044* | *59.7%* | *0.0042* | *34.6%* |
| **Fastest gait speed, m/s** | 14 (799) | 0.10 [0.04, 0.16] | 0.0026 | 0.065 | 62.9% | 0.0008 | - |
| Chronic stroke studies | 10 (617) | 0.07 [0.02, 0.13] | 0.0126 | 0.060 | 59.7% | 0.008 | - |
| Subacute stroke studies | 4 (182) | 0.21 [0.01, 0.41] | 0.046 | 0.060 | 54.8% | 0.0846 | - |
| *Subacute vs chronic stroke* | *14 (799)* | *0.14 [0.01, 0.26]* | *0.0362* | *0.060* | *61.4%* | *0.0019* | *13.3%* |
| **6-minute walk test, m** | 18 (1027) | 37 [29, 46] | <0.0001 | 10.9 | 40.1% | 0.0409 | - |
| Chronic stroke studies | 14 (839) | 33 [24, 42] | <0.0001 | 8.8 | 22.3% | 0.2117 | - |
| Subacute stroke studies | 4 (188) | 51 [26, 77] | 0.0077 | 8.8 | 29.2% | 0.2371 | - |
| *Subacute vs chronic stroke* | *18 (1027)* | *18 [-0, 37]* | *0.0539* | *8.8* | *23.8%* | *0.1781* | *34.4%* |
| **Daily walking activity, steps/day** | 4 (251) | 260 [-1159, 1679] | 0.6008 | 477 | 49.8% | 0.1125 | - |
| For rows with contrasts (rows in *italics*), between-study heterogeneity statistics are for residual heterogeneity not explained by the contrast variable, and R<sup>2</sup> is the % of between-study heterogeneity explained by contrast variable. LT<sub>mv</sub>, moderate-to-vigorous intensity locomotor training; I<sup>2</sup>, % of variance attributed to between-study heterogeneity; SD, standard deviation (between studies); *p-value for between-study heterogeneity is from Cochran's Q test |  |  |  |  |  |  |  |

**Figure 1.**
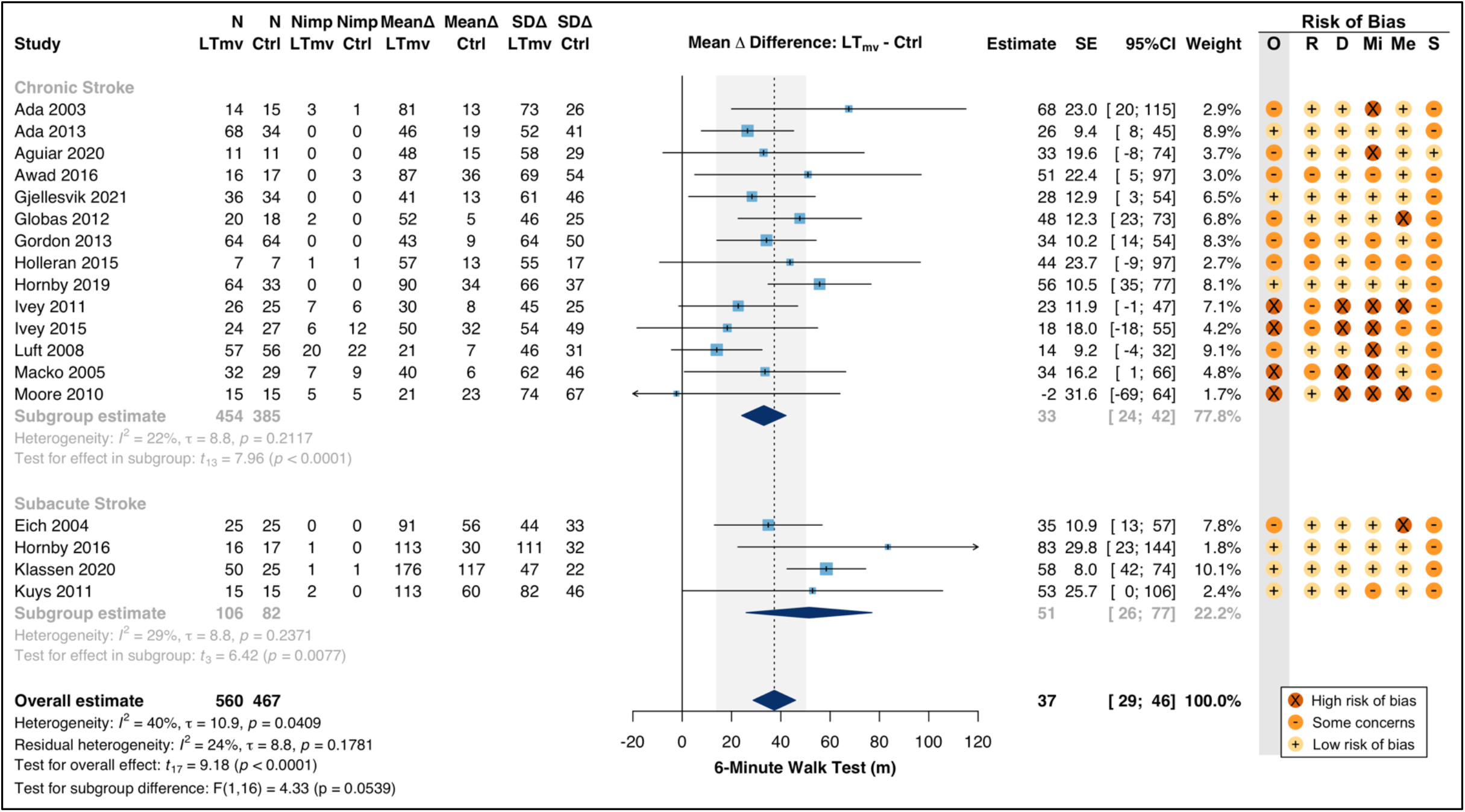
Mean change (Δ) differences for LT_mv_ versus control interventions: 6-minute walk test. Results from a random (mixed) effects meta-analysis stratified by stroke chronicity. The gray shaded region in the forest plot shows a range of clinically important difference thresholds in the literature (14-50 m).^31-33^ Risk-of-bias assessment was performed using the RoB-2 tool, yielding an overall (O) judgment based on the following domains: R, Bias arising from the randomization process; D, Bias due to deviations from intended intervention; Mi, Bias due to missing outcome data; Me, Bias in measurement of the outcome; S, Bias in selection of the reported result. N, number of participants randomized; N_imp_, number of randomized participants not included in the reported analysis whose outcomes were imputed; SD_Δ_, standard deviation of change; LT_mv_, moderate-to-vigorous intensity locomotor training; SE, standard error; Weight, % contribution of each study and subgroup to the overall pooled estimate; I^2^, % of variance attributed to between-study heterogeneity; τ, estimated between-study standard deviation; Heterogeneity p-values are from Cochran’s Q test.

For walking capacity outcomes, study chronicity explained 13-35% of the between-study variance in effect sizes (Table 2). After accounting for chronicity, significant heterogeneity remained for CGS and FGS (with I^2^ of 60-61%) but not the 6MWT (with an I^2^ of 24%). There were insufficient subacute stroke studies to pursue additional meta-regression analyses within that subgroup. In meta-regressions among the chronic stroke studies (Table S1), LT_mv_ mode explained the most between-study variance in CGS and FGS outcomes (54-69%), with significantly greater effect sizes in studies that included some overground gait training versus training only on a treadmill. A similar tendency was observed for 6MWT outcomes (R^2^=30%, p=0.06). Estimated LT_mv_ mean intensity (vigorous versus moderate) explained the most (residual) variance in 6MWT outcomes, but this variable could only be inferred for a subset of studies (11/14) and was not a statistically significant factor (p=0.08). There were insufficient studies to assess the influence of LT_mv_ intensity on CGS or FGS.

### 3.3. LT_mv_ versus control risk of harms

No treatment-related serious adverse effects were reported in any study groups, so relative risks could not be calculated for this effect (Table S2, Fig S5). There were no significant differences in the relative risk of treatment-related pain or the proportion of fallers between LT_mv_ and control groups (Fig S6-S7), and there were insufficient studies for any meta-regressions involving the adverse event measures.

### 3.4. Risk of bias assessments

The subgroup of chronic stroke comparisons judged to be at high risk of bias had significantly *lower* effect sizes for CGS (opposite the typical pattern of concern). Otherwise, there were no clear relationships between risk of bias and outcomes among the included comparisons (Table S1, Fig 1, Figs S2-S7). There was also no statistically significant evidence of publication bias, nor any clear small study asymmetry trends in the funnel plots (Fig S8).

All adverse event comparisons were judged to be at high risk of bias for measurement of the outcome (domain 4). This was driven by more intensive monitoring of LT_mv_ groups in studies where control groups did not include a similar intervention volume (i.e. surveillance bias) and by lack of assessor blinding in all studies, where the study staff assessing adverse events appeared to be the same staff that were delivering the treatment, without any blinded adjudication.

### 3.5. Standard deviation of individual response (SD_IR_)

Sufficient information to calculate SD_IR_ was available in a subset of studies for each outcome (Table 3; Fig 2; Fig S9-S11). The pooled mean SD_IR_ was significantly greater than zero for CGS, almost FGS (p=0.0501) and 6MWT, but not for daily walking activity (p=0.52), which could only pool SD_IR_ data from two studies. There was significant between-study heterogeneity for CGS, FGS and daily walking activity (with I^2^ of 70-86%), but insufficient studies for meta-regression to quantitatively explore its sources. The 6MWT had enough studies for the primary meta-regression but did not have significant heterogeneity (with I^2^ of 27%) and study chronicity did not explain any of the between study variance in SD_IR2_ (R^2^ of 0%).

**Table 3.** Standard Deviations of Individual Response (SD_IR_) to LT_mv_.

| Table 3. Standard Deviations of Individual Response (SD<sub>IR</sub>) to LT<sub>mv</sub> |  |  |  |  |  |  |  |
| --- | --- | --- | --- | --- | --- | --- | --- |
|  |  | Mean Change Difference |  | Between-study heterogeneity |  |  |  |
| | N studies (total N) | Estimate [95%CI] | P-value | SD ($\tau$) | I<sup>2</sup> | P-value* | R<sup>2</sup> |
| **Comfortable gait speed, m/s** | 9 (406) | 0.11 [0.00, 0.15] | 0.0489 | 0.11 | 69.8% | 0.0009 | - |
| **Fastest gait speed, m/s** | 8 (394) | 0.14 [-0.00, 0.20] | 0.0501 | 0.14 | 74.6% | 0.0003 | - |
| **6-minute walk test, m** | 11 (467) | 41 [27, 51] | 0.0025 | 22 | 27.2% | 0.1853 | - |
| Chronic stroke | 8 (357) | 42 [29, 52] | 0.0025 | 25 | 0.0% | 0.4825 | - |
| Subacute stroke | 3 (110) | 38 [-70, 89] | 0.4303 | 25 | 70.3% | 0.0345 | - |
| *Subacute vs chronic stroke* | 11 (467) | -18 [-52, 46] | 0.7707 | 25 | 33.5% | 0.1401 | 0.0% |
| **Daily walking activity (steps/day)** | 2 (90) | 2021 [-7135, 7686] | 0.5185 | 2394 | 86.4% | 0.0067 | - |
| Values were converted from SD<sub>IR</sub><sup>2</sup> units by taking the square root for the estimates, 95% CIs and between-study standard deviations (SD). For the row with a contrast (in *italics*), between-study heterogeneity statistics are for residual heterogeneity not explained by the contrast variable, and R<sup>2</sup> is the % of between-study heterogeneity explained by contrast variable. LT<sub>mv</sub>, moderate-to-vigorous intensity locomotor training; I<sup>2</sup>, % of variance attributed to between-study heterogeneity; *p-value for between-study heterogeneity is from Cochran's Q test |  |  |  |  |  |  |  |

**Figure 2.**
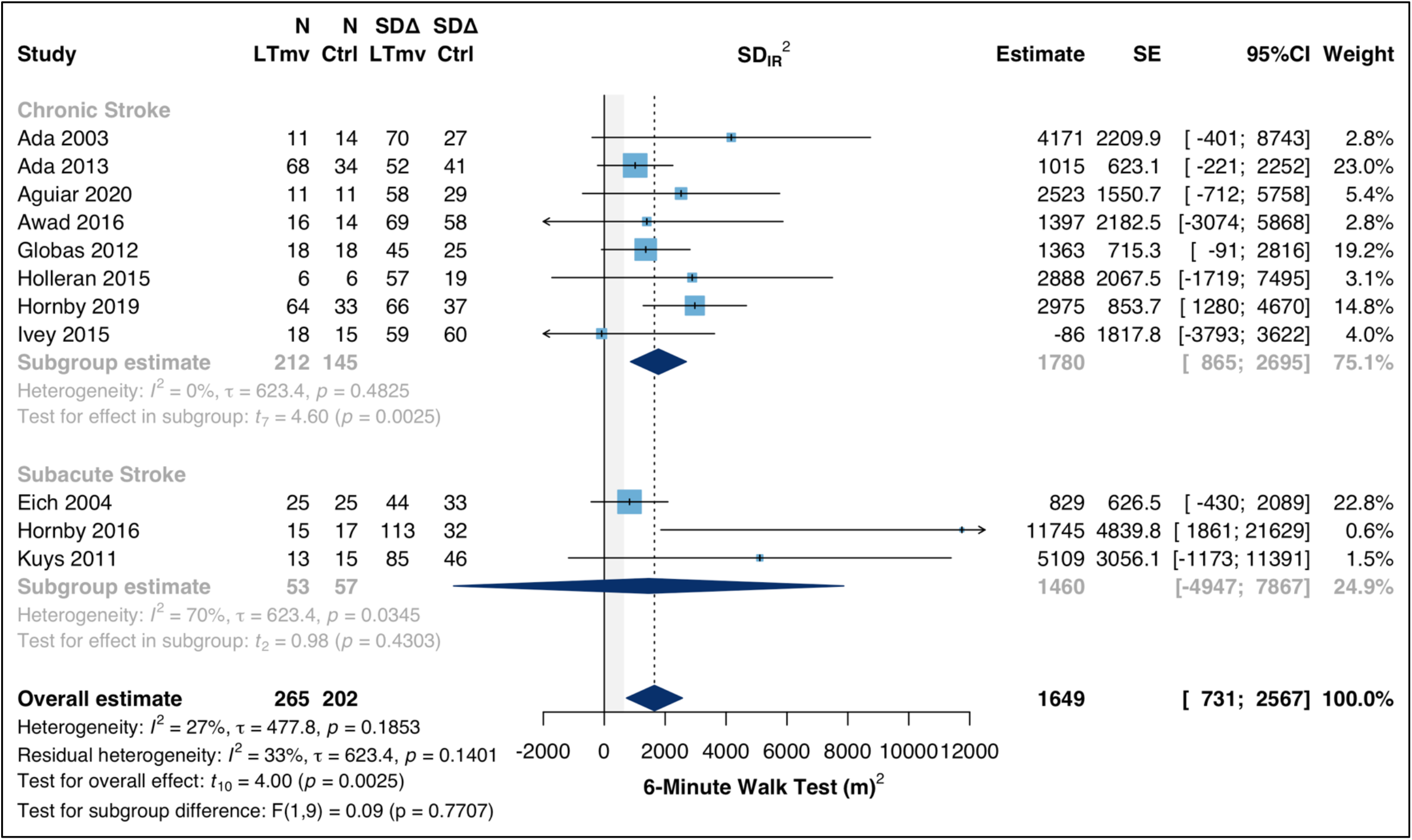
Variance of individual response (SD_IR2_): 6-minute walk test. Results from a random (mixed) effects meta-analysis stratified by stroke chronicity. SD_IR_ estimates and 95%CI can be obtained by taking the square root of the values in this figure. The gray shaded region in the forest plot shows a range of clinically important difference thresholds in the literature (14-50 m),^31-33^ converted to SD_IR2_ units by halving and squaring (49-625 m^2^).^4,16^ N, number of participants analyzed; SDΔ, standard deviation of change; LT_mv_, moderate-to-vigorous intensity locomotor training; SE, standard error; Weight, % contribution of each study and subgroup to the overall pooled estimate; I^2^, % of variance attributed to between-study heterogeneity; τ, estimated between-study standard deviation; Heterogeneity p-values are from Cochran’s Q test.

### 3.6. Estimated proportions of responders

The estimated proportion of participants with at least small net changes attributable to LT_mv_ ranged across capacity measures from 36-68% in chronic stroke and from 71-82% in subacute stroke (Fig 3).

**Figure 3.**
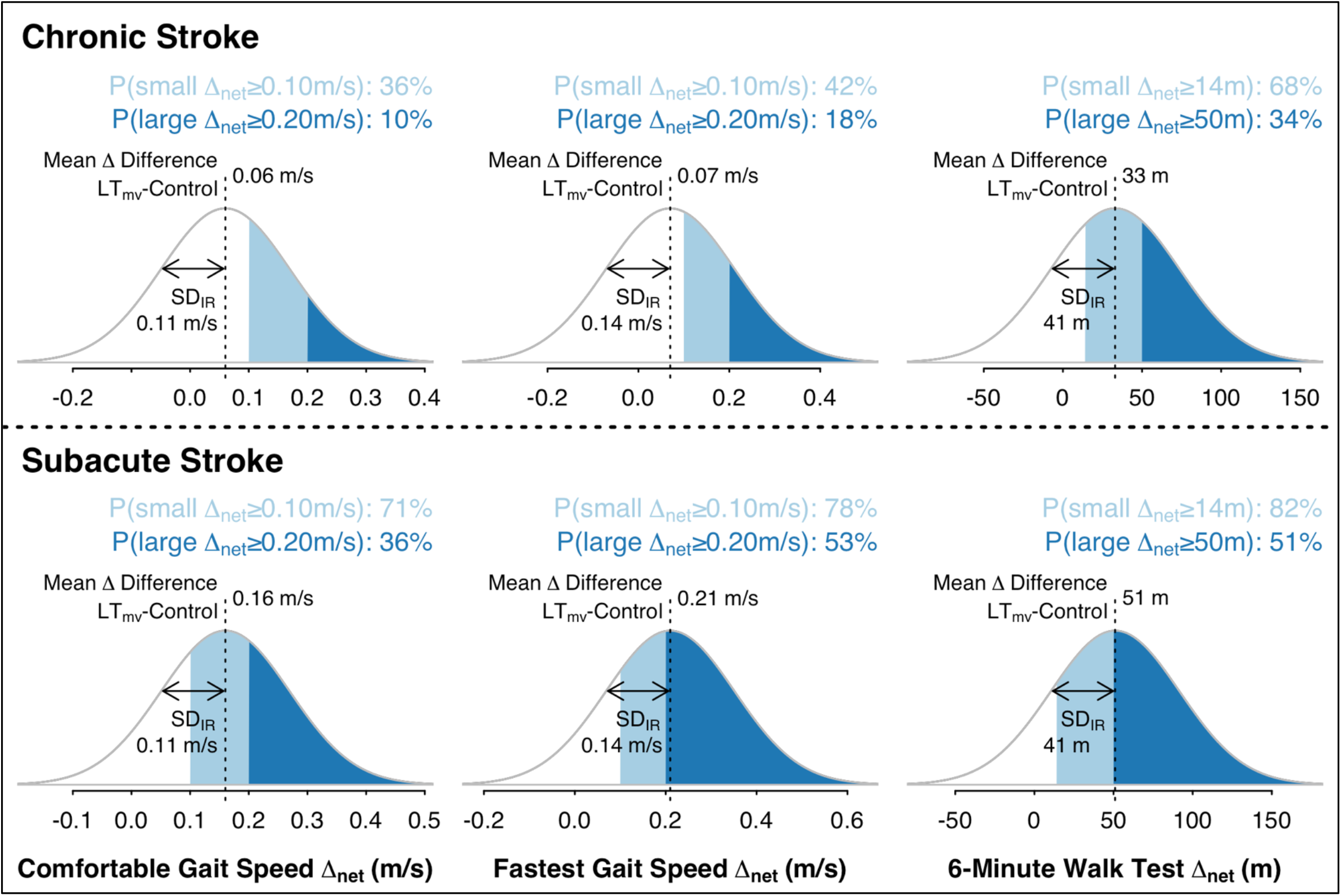
Estimated proportions of participants with meaningful gains attributable to LT_mv_. Graphs show the estimated distributions of individual differences in response (ι1_net_) to LT_mv_ versus control (i.e. how much more [or less] individuals are expected to improve with LT_mv_ relative to control interventions), based on the meta-analysis results. Separate distributions were calculated for each outcome measure (from left to right: comfortable gait speed, fastest gait speed, 6-minute walk test) and for each stroke chronicity subgroup (top panel, chronic; bottom panel, subacute). The mean of each distribution is the meta-analysis estimate of the mean ι1 difference (LT_mv_ – control), and the SD of each distribution is the meta-analysis estimate of the SD_IR_ (the estimate of the true individual variability in responsiveness).^4,14^ Reported probabilities (P) are the % of individual net changes that were at or above different clinically important difference thresholds from the literature. These thresholds ranged from 0.10 m/s (small) to 0.20 m/s (large) for gait speed^28-31^ and from 14 m (small) to 50 m (large) for the 6-minute walk test.^31-33^

Estimated proportions of responders with large net changes were 10-34% in chronic stroke and 36-53% in subacute stroke. Pooled estimates of mean changes within LT_mv_ groups were also calculated to facilitate clinical interpretation (Table S3, Figs S12-S15). Based on these values, the estimated proportions of participants with at least small changes during LT_mv_ (not necessarily attributable to LT_mv_) were 54-81% in chronic stroke and 99-100% in subacute stroke (Fig S16). Estimated proportions of responders with large changes were 21-50% in chronic stroke and 93-97% in subacute stroke.

### 4. DISCUSSION

This meta-analysis synthesized data on outcomes, risk of harms and response variability for LT_mv_ in subacute and chronic stroke recovery. Compared with no intervention, non-gait intervention or low-intensity gait training controls, LT_mv_ yielded significantly greater improvement in all walking capacity outcomes (CGS, FGS and 6MWT) in both subacute and chronic stroke, with subacute studies showing greater effect sizes (Table 2). Based on the smallest CID threshold for these outcomes (0.10 m/s for CGS/FGS^28-31^ and 14 m for 6MWT^31-33^), mean LT_mv_ gains were meaningfully better than control groups for the 6MWT in chronic stroke studies, and for all three walking capacity outcomes in subacute studies. These positive outcomes did not appear to be driven by individual study bias or publication bias. There were also no serious adverse effects related to treatment among 398 LT_mv_ participants, suggesting <0.25% risk of serious harms. These findings strengthen the recent clinical practice guideline recommendation that LT_mv_ should be used in chronic stroke,^1^ and appear to warrant extending this recommendation to subacute stroke.

Nevertheless, SD_IR_ estimates indicated significant individual variability in LT_mv_ response for CGS, 6MWT and nearly FGS (p=0.0501). Each of these SD_IR_ estimates exceeded all CID thresholds (in SD units:^4,16^ 0.05-0.10 m/s for CGS/FGS^28-31^ and 7-25 m for 6MWT^31-33^), signifying a large and meaningful amount of individual response variability. Thus, clinicians and patients/clients should be aware that group-level LT_mv_ outcomes may not be very informative for expected individual changes. Future studies to improve prediction of individual LT_mv_ responses are strongly indicated.

Based on the mean change differences and SD_IR_ results, we estimated that LT_mv_ would elicit at least small meaningful gains in walking capacity outcomes (that would not otherwise occur with control interventions) for 36-68% of chronic stroke survivors and 71-82% of subacute stroke survivors (Fig 3). These estimates imply that LT_mv_ remains a powerful intervention despite response variability, producing meaningful walking capacity gains in 1 out of every 1-3 chronic stroke survivors treated and 1 out of every 1-2 subacute stroke survivors treated. When ignoring the control groups to mimic LT_mv_ delivery in clinical practice, we estimated that meaningful gains in walking capacity outcomes *during* LT_mv_ (but not necessarily attributable to LT_mv_) would occur for 54-81% of chronic stroke survivors and 99-100% of subacute stroke survivors (Fig S16), at least in patients/clients similar to the included study samples. These percentages provide useful information for managing expectations in clinical practice.

While our findings indicate that LT_mv_ is efficacious for improving walking *capacity*, its effects on daily walking activity are less clear, as only four (out of 19) studies contributed to the pooled estimate of LT_mv_ versus control changes in steps/day (+260 [95%CI: -1,159 to 1,679]). In the absence of published CID thresholds for steps/day post-stroke, we interpret the point estimate of 260 steps/day as a small non-meaningful difference, but interpret the 95%CI bounds as very large differences. Thus, our current best estimate suggests LT_mv_ may not have a meaningful effect on daily walking activity, but this estimate is imprecise, and we cannot rule out very large beneficial or detrimental effects.

A lack of meaningful change in walking activity with LT_mv_ would be consistent with recent findings that gains in walking capacity do not automatically translate into gains in daily walking activity post-stroke.^52,53^ This limited translation may be due to the complex, multifactorial nature of daily walking activity, which requires sufficient walking capacity but can also be influenced by many other personal and environmental factors.^54-57^ The multifactorial nature of this measure may also explain the extremely large (albeit non-significant) individual variability we observed in walking activity changes from LT_mv_ (pooled SD_IR_: 2,021 steps/day [95% CI: -7,135, 7,686]). Further studies assessing daily walking activity are needed to develop its CID benchmarks, more precisely estimate LT_mv_ effects, and augment the effects of LT_mv_ on this outcome.

Moderate between-study heterogeneity was present in the overall meta-analyses for all outcomes (I^2^, 40-68%; Table 2), matching our expectation that the magnitude of LT_mv_ effects likely depends on some study characteristics (e.g. participant sample, LT_mv_ delivery methods). Stroke chronicity explained 13-35% of this heterogeneity (across walking capacity outcomes) and there were enough chronic stroke studies to test how well other study factors explained residual heterogeneity within this subgroup (Table S1). In those analyses, studies that included some overground gait training had significantly greater improvements in CGS, FGS and nearly 6MWT (p=0.06), compared with studies involving only treadmill training. Like previous research,^58^ this suggests that optimal LT_mv_ delivery may include both treadmill and overground training. Our results also hinted that vigorous LT_mv_ intensity may be superior to moderate intensity (like other research^59^), but training intensity was difficult to determine from many of the reports, could only be tested for its association with 6MWT effect sizes and was not a significant factor (p=0.08).

Adverse event reporting was generally unsystematic across the included studies, and mostly appeared to reflect passive surveillance (i.e. relying on participants to self-report) without any specific queries or monitoring for events of interest. For example, several studies only reported that there were no adverse events, without specifying what types of events would have been reported had they occurred. This was less problematic for assessing serious adverse effects related to treatment, because it seemed safe to assume those would have been observed and reported if present, at least in the 14 (out of 19) studies that included some mention of adverse events. Thus, we are more confident in our estimate that <1 in 398 participants have serious adverse effects from LT_mv_, and less confident in our relative risk estimates for non-serious treatment-related pain (1.91 [95%CI: 0.80, 4.54]) and non-serious falls outside of treatment (0.89 [95%CI: 0.57, 1.37]).

While imprecise and not statistically significant, these current point estimates imply that the risk of treatment-related pain (e.g. muscle/joint soreness) could be 91% higher with LT_mv_ versus control interventions (including no treatment), and that fall risk could be reduced by 11%. If accurate, these relative risks suggest that LT_mv_ could cause mild/moderate pain (that would not otherwise occur with control interventions) in 1 out of every 10 stroke survivors treated and could prevent falls in 1 out of every 35 (calculations^2^ based on control group event rates in Table S2). From these non-serious event rates and the numbers needed to treat for meaningful walking capacity gains above, LT_mv_ benefits post-stroke appear to strongly outweigh its risks on average. Still, adverse event comparisons from all included studies were judged to have at least some concerns of overall bias, and high risk of bias for the outcome measurement domain (Figs S5-S7). This was primarily driven by unblinded adverse event assessment by the same personnel delivering treatment without any blinded adjudication. Thus, future LT_mv_ studies are needed to assess adverse events more systematically with larger samples and less risk of bias.

### 4.1. Strengths and Limitations

Methodological strengths of this meta-analysis include: the use of random effects models to account for plausible between-study heterogeneity and meta-regression to partially explain it; pooling outcomes using the original measurement scale (i.e. unstandardized effect sizes) for better clinical interpretability than using standard deviation units; pooling data on adverse events to better elucidate the balance between risk and benefit; thorough assessment of risk of bias; pooled estimation of individual response variability in addition to mean effects; novel accounting for between-group differences in variance for more appropriate estimation of standard errors within individual studies; missing data imputation to estimate intent-to-treat effects for each study when not reported by the authors; and estimating proportions of responders to enhance interpretability of the results.

One limitation of this meta-analysis is that it did not account for the uncertainty associated with the estimation of between study variance (e.g. using Bayesian methods^22^). Several additional limitations apply to the meta-regression analyses. Since meta-regression is observational, confounding can produce non-causal associations and can obscure causal effects. We used a principled approach to decrease the impact of confounding (e.g. only testing models with a reasonable number of studies, accounting for stroke chronicity in all models), but there were insufficient studies to thoroughly assess or control for other plausible confounders. Another meta-regression limitation is that many of the independent variables tested have variability at the individual level, but could only be extracted at the group level, which can produce misleading results (i.e. ecologic fallacy^18^). In addition, some meta-regression variables were difficult to ascertain for individual studies and required inference during data extraction.

## 5. Conclusions

LT_mv_ improves mean walking capacity outcomes in both subacute and chronic stroke and does not appear to have high risk of serious harm, but response magnitude varies between chronicity subgroups and individuals, and few studies have tested effects on daily walking activity or non-serious adverse events.

## Data Availability

All data produced in the present work are contained in the manuscript

https://sysrev.com/u/5631/p/93708

https://sysrev.com/u/5631/p/103118

---

**Table S1.** Meta-Regressions for LT_mv_-Control Mean Change Differences Within Chronic Stroke Subgroup.

|  |  | Mean Change Difference |  | Between-study heterogeneity |  |  |  |
| --- | --- | --- | --- | --- | --- | --- | --- |
| Within Chronic Stroke Studies | N studies (total N) | Estimate [95%CI] | P-value | SD ($\tau$) | I<sup>2</sup> | P-value* | R<sup>2</sup> |
| **Comfortable gait speed (CGS), m/s** | 10 (514) | 0.06 [0.01, 0.10] | 0.0173 | 0.044 | 65.1% | 0.0022 | - |
| *Mean baseline CGS (each 0.1 m/s ↑)* | 10 (514) | -0.01 [-0.04, 0.03] | 0.5588 | 0.053 | 69.9% | 0.0008 | 0.0% |
| Treadmill-only LT<sub>mv</sub> | 6 (253) | 0.02 [-0.03, 0.07] | 0.3826 | 0.028 | 30.1% | 0.2092 | - |
| Some overground LT<sub>mv</sub> | 4 (261) | 0.11 [0.06, 0.16] | 0.0068 | 0.028 | 7.0% | 0.3579 | - |
| *Some overground LT<sub>mv</sub> vs treadmill only* | 10 (514) | 0.09 [0.03, 0.15] | 0.0100 | 0.028 | 23.9% | 0.2305 | 68.5% |
| *Planned LT<sub>mv</sub> volume (each 10 hour ↑)* | 10 (514) | -0.02 [-0.06, 0.01] | 0.2156 | 0.043 | 60.1% | 0.0101 | 23.5% |
| Control with no gait training | 5 (250) | 0.07 [0.03, 0.11] | 0.0093 | 0.054 | 0.0% | 0.7988 | - |
| Control with some gait training | 5 (264) | 0.04 [-0.06, 0.15] | 0.3266 | 0.054 | 83.4% | <0.0001 | - |
| *Control with vs without gait training* | 10 (514) | -0.03 [-0.12, 0.07] | 0.5432 | 0.054 | 72.1% | 0.0004 | 0.0% |
| Low/some risk of bias | 6 (321) | 0.09 [0.03, 0.14] | 0.0086 | 0.032 | 49.3% | 0.0794 | - |
| High risk of bias | 4 (193) | 0.01 [-0.07, 0.08] | 0.7612 | 0.032 | 16.3% | 0.3098 | - |
| *High vs low/some risk of bias* | 10 (514) | -0.08 [-0.16, -0.00] | 0.0423 | 0.032 | 42.0% | 0.0875 | 58.9% |
| **Fastest gait speed (FGS), m/s** | 10 (617) | 0.07 [0.02, 0.13] | 0.0126 | 0.060 | 59.7% | 0.008 | - |
| *Mean baseline FGS (each 0.1 m/s ↑)* | 10 (617) | -0.01 [-0.02, 0.01] | 0.2759 | 0.058 | 61.8% | 0.0073 | 3.0% |
| Treadmill-only LT<sub>mv</sub> | 7 (385) | 0.04 [-0.01, 0.10] | 0.1126 | 0.040 | 24.7% | 0.2407 | - |
| Some overground LT<sub>mv</sub> | 3 (232) | 0.14 [-0.00, 0.28] | 0.0505 | 0.040 | 50.1% | 0.1350 | - |
| *Some overground LT<sub>mv</sub> vs treadmill only* | 10 (617) | 0.10 [0.01, 0.19] | 0.0312 | 0.040 | 34.7% | 0.1405 | 54.2% |
| *Planned LT<sub>mv</sub> volume (each 10 hour ↑)* | 10 (617) | -0.03 [-0.07, 0.01] | 0.1434 | 0.046 | 48.8% | 0.0483 | 39.9% |
| Control with no gait training | 5 (353) | 0.08 [0.02, 0.14] | 0.0167 | 0.065 | 2.3% | 0.3936 | - |
| Control with some gait training | 5 (264) | 0.06 [-0.06, 0.19] | 0.2396 | 0.065 | 78.0% | 0.0011 | - |
| *Control with vs without gait training* | 10 (617) | -0.02 [-0.13, 0.10] | 0.7306 | 0.065 | 67.5% | 0.0018 | 0.0% |
| Low/some risk of bias | 7 (475) | 0.10 [0.02, 0.17] | 0.0183 | 0.051 | 63.2% | 0.0123 | - |
| High risk of bias | 3 (142) | 0.02 [-0.03, 0.07] | 0.2616 | 0.051 | 0.0% | 0.9176 | - |
| *High vs low/some risk of bias* | 10 (617) | -0.08 [-0.20, 0.05] | 0.1889 | 0.051 | 53.9% | 0.0266 | 26.2% |
| **6-minute walk test (6MWT), m** | 14 (839) | 33 [24, 42] | <0.0001 | 8.8 | 22.3% | 0.2117 | - |
| *Mean baseline 6MWT (each 10 m ↑)* | 14 (839) | 0 [-1, 1] | 0.8349 | 9.4 | 28.1% | 0.1612 | 0.0% |
| Treadmill-only LT<sub>mv</sub> | 8 (436) | 26 [15, 37] | 0.0008 | 7.1 | 0.0% | 0.5116 | - |
| Some overground LT<sub>mv</sub> | 6 (403) | 41 [26, 57] | 0.0010 | 7.1 | 20.7% | 0.2775 | - |
| *Some overground LT<sub>mv</sub> vs treadmill only* | 14 (839) | 15 [-1, 32] | 0.0622 | 7.1 | 4.7% | 0.399 | 29.6% |
| Moderate LT<sub>mv</sub> intensity | 6 (397) | 25 [12, 37] | 0.0036 | 4.9 | 0.0% | 0.5609 | - |
| Vigorous LT<sub>mv</sub> intensity | 5 (278) | 41 [22, 60] | 0.0038 | 4.9 | 19.9% | 0.2876 | - |
| *Vigorous vs moderate intensity* | 11 (675) | 16 [-2, 34] | 0.0791 | 4.9 | 0.0% | 0.444 | 71.6% |
| *Planned LT<sub>mv</sub> volume (each 10 hour ↑)* | 14 (839) | -6 [-11, 0] | 0.0600 | 4.9 | 0.6% | 0.4395 | 66.0% |
| Control with no gait training | 8 (561) | 29 [17, 41] | 0.0008 | 5.5 | 25.1% | 0.2286 | - |
| Control with some gait training | 6 (278) | 42 [26, 58] | 0.0009 | 5.5 | 0.0% | 0.5378 | - |
| *Control with vs without gait training* | 14 (839) | 13 [-5, 32] | 0.1431 | 5.5 | 10.8% | 0.3371 | 58.1% |
| Low/some risk of bias | 10 (646) | 36 [25, 47] | <0.0001 | 8.9 | 35.3% | 0.1259 | - |
| High risk of bias | 4 (193) | 22 [5, 40] | 0.0261 | 8.9 | 0.0% | 0.7676 | - |
| *High vs low/some risk of bias* | 14 (839) | -14 [-35, 7] | 0.1835 | 8.9 | 20.9% | 0.2323 | 0.0% |
| For rows with contrasts (rows in *italics*), between-study heterogeneity statistics are for residual heterogeneity not explained by the contrast variable, and R<sup>2</sup> is the % of between-study heterogeneity explained by contrast variable. LT<sub>mv</sub>, moderate-to-vigorous intensity locomotor training; I<sup>2</sup>, % of variance attributed to between-study heterogeneity; SD, standard deviation; *p-value for between-study heterogeneity is from Cochran's Q test; |  |  |  |  |  |  |  |

**Table S2.** LT_mv_ Versus Control Risks of Harm.

| **Table S2. LT<sub>mv</sub> Versus Control Risks of Harm** |  |  |  |  |  |  |  |  |  |
| --- | --- | --- | --- | --- | --- | --- | --- | --- | --- |
|  | **N studies**<br>**(total N)**<br>**reporting**<br>**on event** | **N events / N**<br>**participants (%)** |  | **Relative Risk**<br>**(LT<sub>mv</sub> / Control)** |  |  | **Between-study**<br>**heterogeneity** |  |  |
|  |  | **LT<sub>mv</sub>** | **Control** | **N studies**<br>**(total N)\*** | **Estimate**<br>**[95%CI]** | **P-**<br>**value** | **SD**<br>**(τ)** | **I<sup>2</sup>** | **P-**<br>**value\*** |
| **Treatment-Related SAE** | 14 (758) | 0 / 398<br>(0.0%) | 0 / 360<br>(0.0%) | 0 (0) | - | - | - | - | - |
| **Treatment-Related Pain** | 7 (350) | 56 / 182<br>(30.8%) | 19 / 168<br>(11.3%) | 4 (190) | 1.91 [0.80, 4.54] | 0.1433 | 0.49 | 0% | 0.4060 |
| **Falls** | 6 (287) | 43 / 172<br>(25.0%) | 30 / 115<br>(26.1%) | 5 (269) | 0.89 [0.57, 1.37] | 0.5861 | 0.21 | 12% | 0.3365 |
| *Studies with no events in either group do not contribute any information about relative risks between groups. LT<sub>mv</sub>, moderate-to-vigorous intensity locomotor training; I<sup>2</sup>, % of variance attributed to between-study heterogeneity; *p-value for between-study heterogeneity is from Cochran's Q test; SAE, serious adverse effect; |  |  |  |  |  |  |  |  |  |

**Table S3.** Mean Changes Within LT_mv_ Groups.

|  |  | **Mean Change Within LT<sub>mv</sub> Groups** |  | **Between-study heterogeneity** |  |  |  |
| --- | --- | --- | --- | --- | --- | --- | --- |
|  | **N studies (total N)** | **Estimate [95%CI]** | **P-value** | **SD ($\tau$)** | **I<sup>2</sup>** | **P-value\*** | **R<sup>2</sup>** |
| **Comfortable gait speed, m/s** | 13 (652) | 0.16 [0.08, 0.24] | 0.0008 | 0.126 | 96.9% | <0.0001 | - |
| Chronic stroke studies | 10 (514) | 0.11 [0.06, 0.15] | 0.0003 | 0.073 | 77.0% | <0.0001 | - |
| Subacute stroke studies | 3 (138) | 0.36 [-0.00, 0.73] | 0.0512 | 0.073 | 93.3% | <0.0001 | - |
| *Subacute vs chronic stroke* | 13 (652) | 0.25 [0.13, 0.37] | 0.0007 | 0.073 | 89.0% | <0.0001 | 66.4% |
| **Fastest gait speed, m/s** | 14 (799) | 0.20 [0.08, 0.33] | 0.004 | 0.196 | 90.4% | <0.0001 | - |
| Chronic stroke studies | 10 (617) | 0.12 [0.06, 0.18] | 0.0018 | 0.122 | 82.3% | <0.0001 | - |
| Subacute stroke studies | 4 (182) | 0.43 [-0.02, 0.87] | 0.0546 | 0.122 | 89.4% | <0.0001 | - |
| *Subacute vs chronic stroke* | 14 (799) | 0.31 [0.11, 0.52] | 0.0057 | 0.122 | 86.5% | <0.0001 | 61.6% |
| **6-minute walk test, m** | 18 (1027) | 66 [46, 86] | <0.0001 | 38.3 | 95.5% | <0.0001 | - |
| Chronic stroke studies | 14 (839) | 50 [37, 62] | <0.0001 | 23.8 | 74.7% | <0.0001 | - |
| Subacute stroke studies | 4 (188) | 127 [61, 193] | 0.0088 | 23.8 | 95.3% | <0.0001 | - |
| *Subacute vs chronic stroke* | 18 (1027) | 77 [45, 110] | 0.0001 | 23.8 | 87.1% | <0.0001 | 61.5% |
| **Daily walking activity, steps/day** | 4 (251) | 270 [-1518, 2057] | 0.664 | 937 | 72.2% | 0.0129 | - |
| For rows with contrasts (rows in *italics*), between-study heterogeneity statistics are for residual heterogeneity not explained by the contrast variable, and R<sup>2</sup> is the % of between-study heterogeneity explained by contrast variable. LT<sub>mv</sub>, moderate-to-vigorous intensity locomotor training; I<sup>2</sup>, % of variance attributed to between-study heterogeneity; SD, standard deviation; *p-value for between-study heterogeneity is from Cochran's Q test |  |  |  |  |  |  |  |

**Figure S1.**
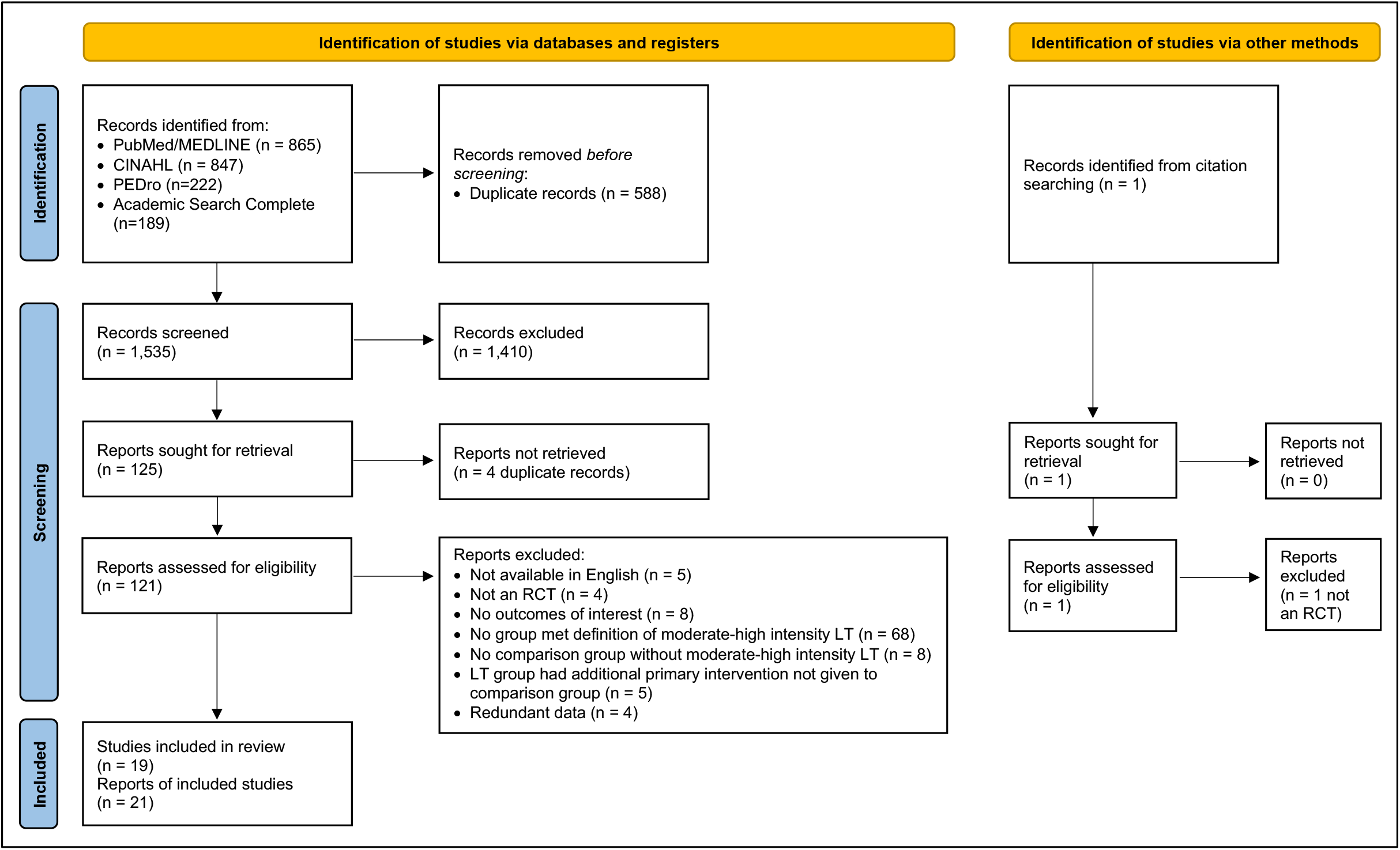
PRISMA Flow Diagram.

**Figure S2.**
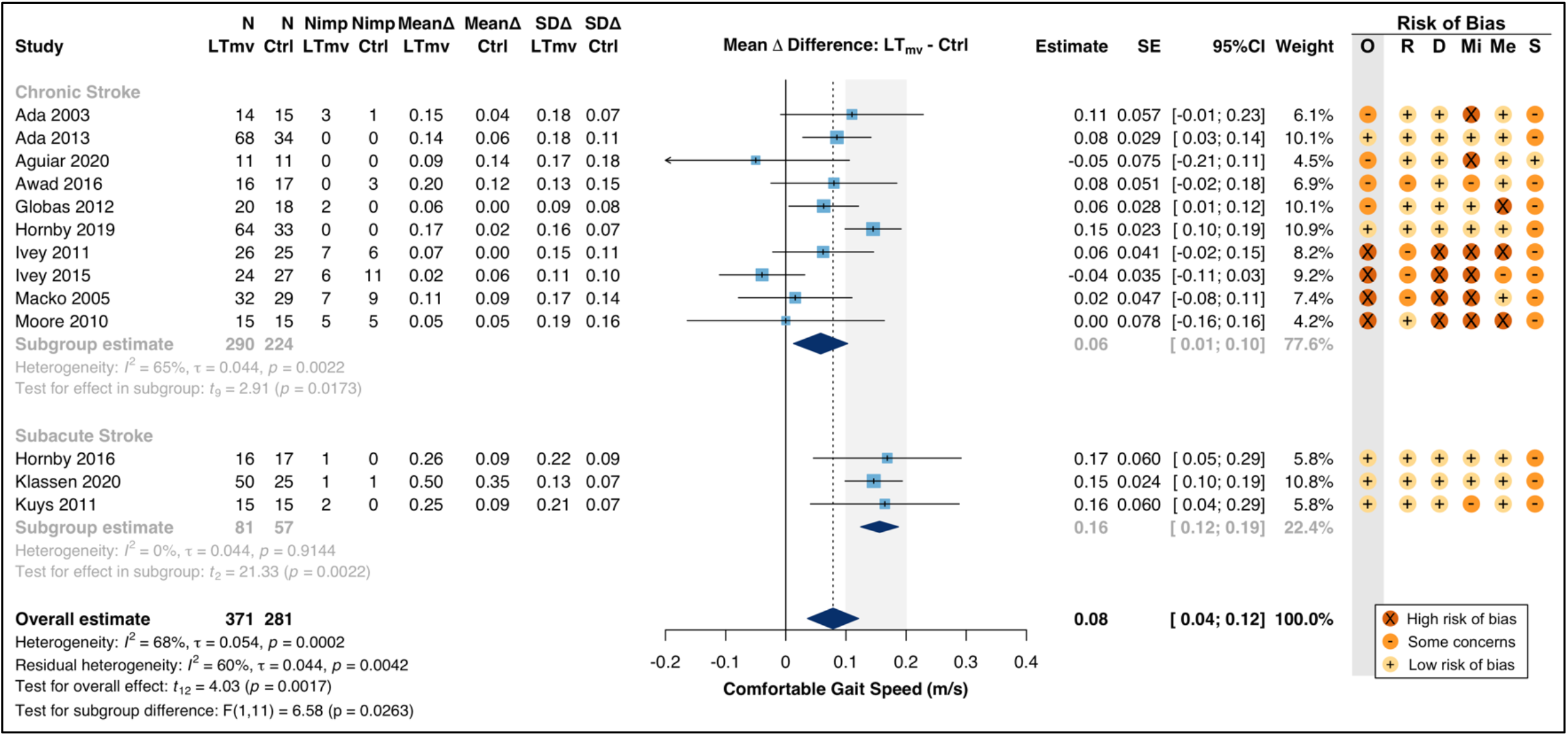
Mean change (Δ) differences for LT_mv_ versus control interventions: Comfortable gait speed. Results from a random (mixed) effects meta-analysis stratified by stroke chronicity. The gray shaded region in the forest plot shows a range of clinically important difference thresholds in the literature (0.1-0.2 m/s). Risk-of-bias assessment was performed using the RoB-2 tool, yielding an overall (O) judgment based on the following domains: R, Bias arising from the randomization process; D, Bias due to deviations from intended intervention; Mi, Bias due to missing outcome data; Me, Bias in measurement of the outcome; S, Bias in selection of the reported result. N, number of participants randomized; N_imp_, number of randomized participants not included in the reported analysis whose outcomes were imputed; SDΔ, standard deviation of change; LT_mv_, moderate-to-vigorous intensity locomotor training; SE, standard error; Weight, % contribution of each study and subgroup to the overall pooled estimate; I^2^, % of variance attributed to between-study heterogeneity; τ, estimated between-study standard deviation; Heterogeneity p-values are from Cochran’s Q test.

**Figure S3.**
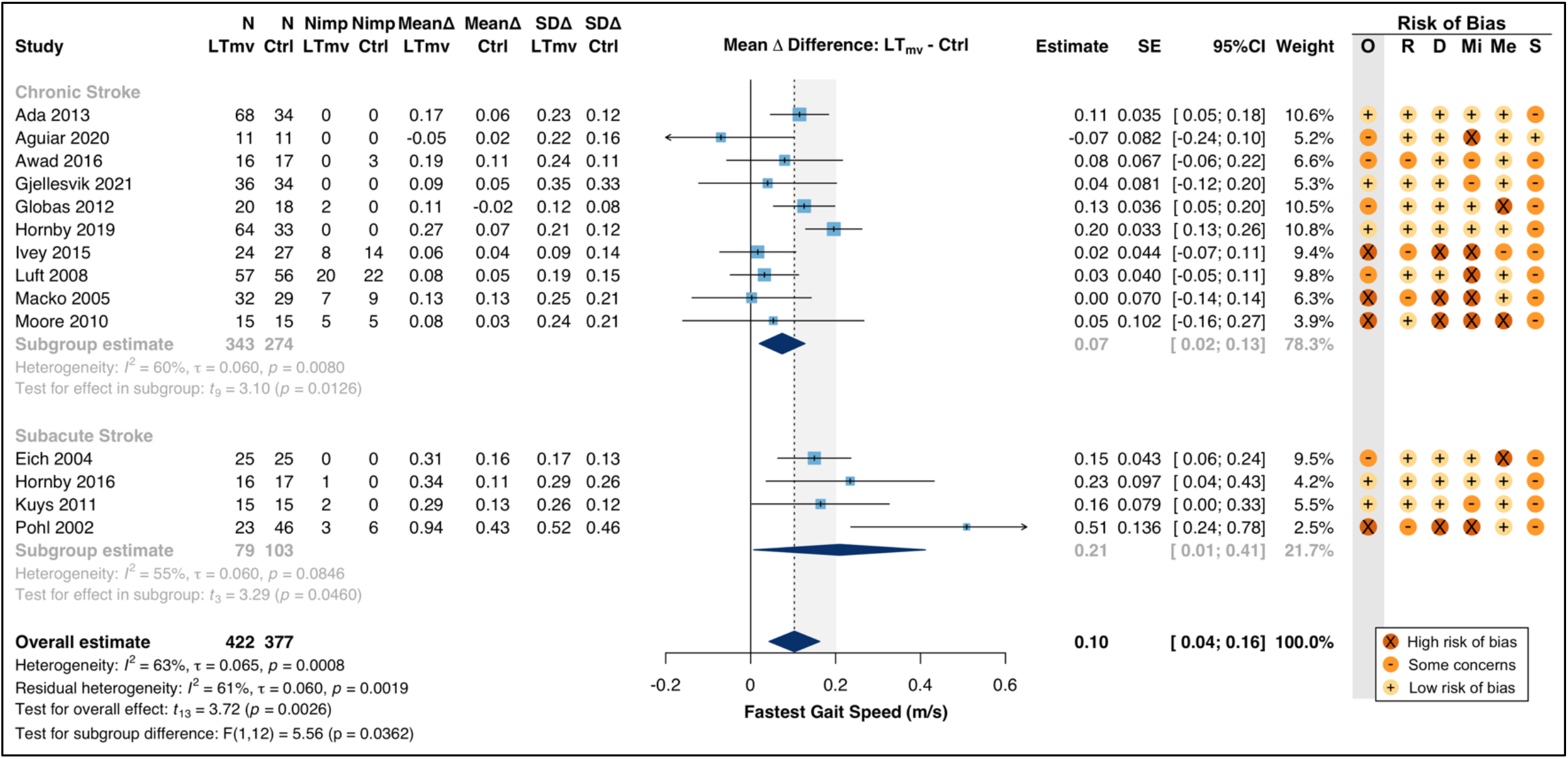
Mean change (Δ) differences for LT_mv_ versus control interventions: Fastest gait speed. Results from a random (mixed) effects meta-analysis stratified by stroke chronicity. The gray shaded region in the forest plot shows a range of clinically important difference thresholds in the literature (0.1-0.2 m/s). Risk-of-bias assessment was performed using the RoB-2 tool, yielding an overall (O) judgment based on the following domains: R, Bias arising from the randomization process; D, Bias due to deviations from intended intervention; Mi, Bias due to missing outcome data; Me, Bias in measurement of the outcome; S, Bias in selection of the reported result. N, number of participants randomized; N_imp_, number of randomized participants not included in the reported analysis whose outcomes were imputed; SDΔ, standard deviation of change; LT_mv_, moderate-to-vigorous intensity locomotor training; SE, standard error; Weight, % contribution of each study and subgroup to the overall pooled estimate; I^2^, % of variance attributed to between-study heterogeneity; τ, estimated between-study standard deviation; Heterogeneity p-values are from Cochran’s Q test.

**Figure S4.**
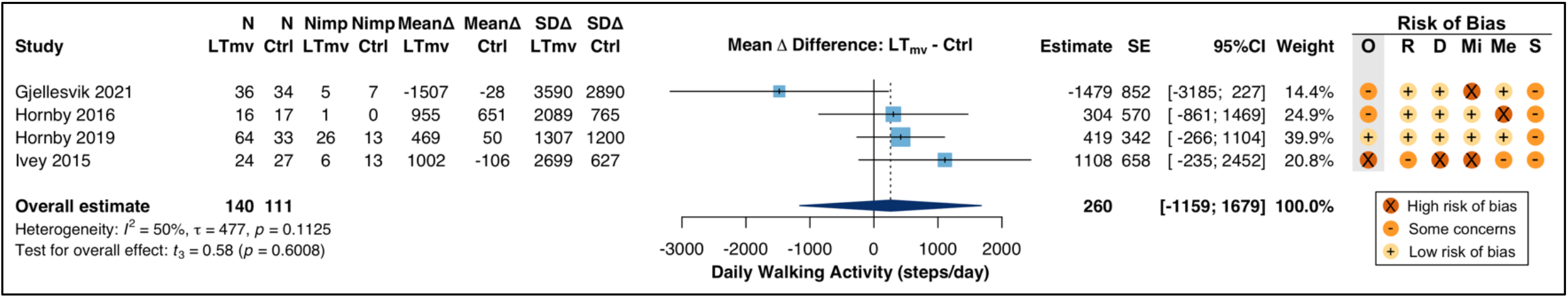
Mean change (Δ) differences for LT_mv_ versus control interventions: Daily walking activity. Results from a random effects meta-analysis. Risk-of-bias assessment was performed using the RoB-2 tool, yielding an overall (O) judgment based on the following domains: R, Bias arising from the randomization process; D, Bias due to deviations from intended intervention; Mi, Bias due to missing outcome data; Me, Bias in measurement of the outcome; S, Bias in selection of the reported result. N, number of participants randomized; N_imp_, number of randomized participants not included in the reported analysis whose outcomes were imputed; SDΔ, standard deviation of change; LT_mv_, moderate-to-vigorous intensity locomotor training; SE, standard error; Weight, % contribution of each study to the overall pooled estimate; I^2^, % of variance attributed to between-study heterogeneity; τ, estimated between-study standard deviation; Heterogeneity p-values are from Cochran’s Q test.

**Figure S5.**
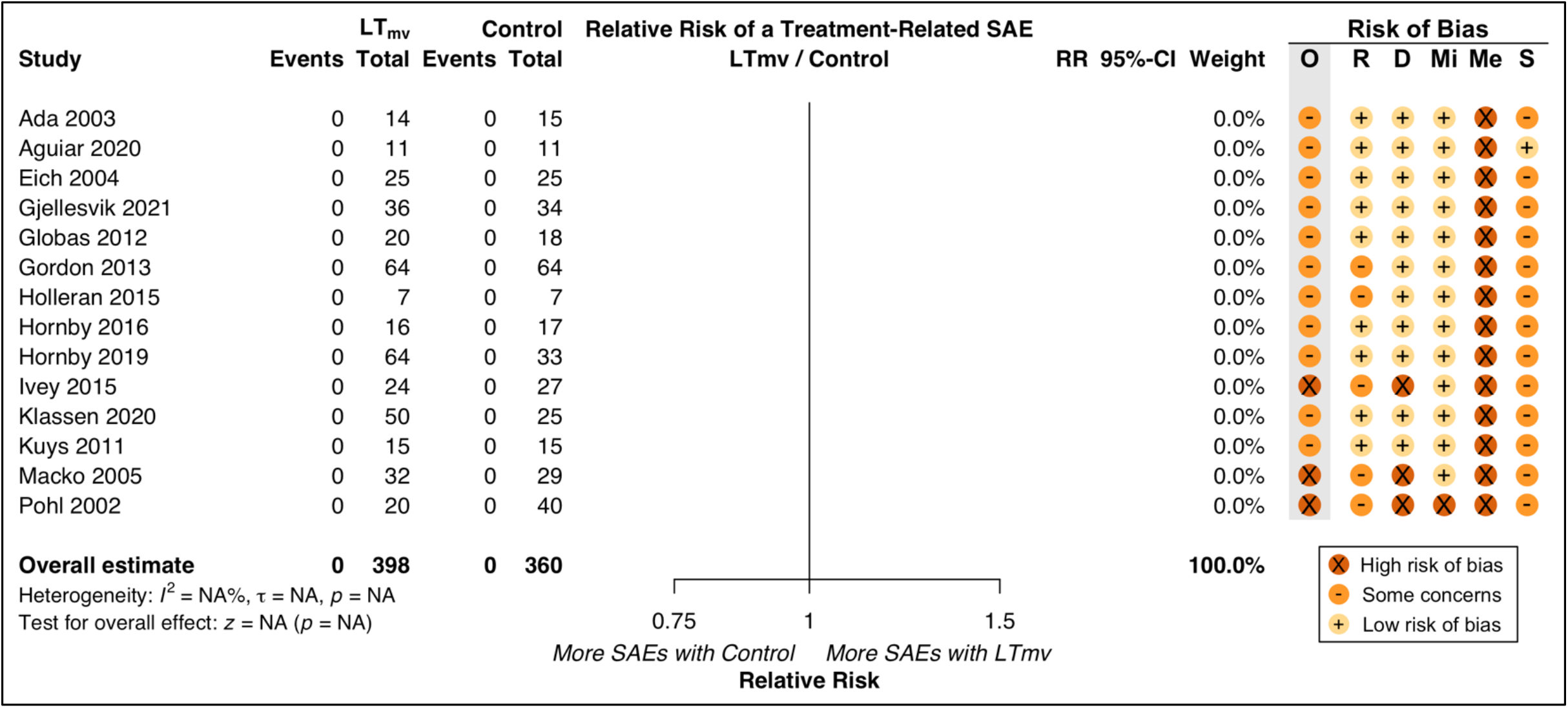
Relative risk (RR) of a treatment-related serious adverse effect (SAE) for LT_mv_ versus control interventions. Unable to perform meta-analysis because no treatment-related SAEs were reported in any study group. Only studies with some reporting about adverse events are shown. Risk-of-bias assessment was performed using the RoB-2 tool, yielding an overall (O) judgment based on the following domains: R, Bias arising from the randomization process; D, Bias due to deviations from intended intervention; Mi, Bias due to missing outcome data; Me, Bias in measurement of the outcome; S, Bias in selection of the reported result. LT_mv_, moderate-to-vigorous intensity locomotor training; Total, number of participants exposed to at least some of the intervention.

**Figure S6.**
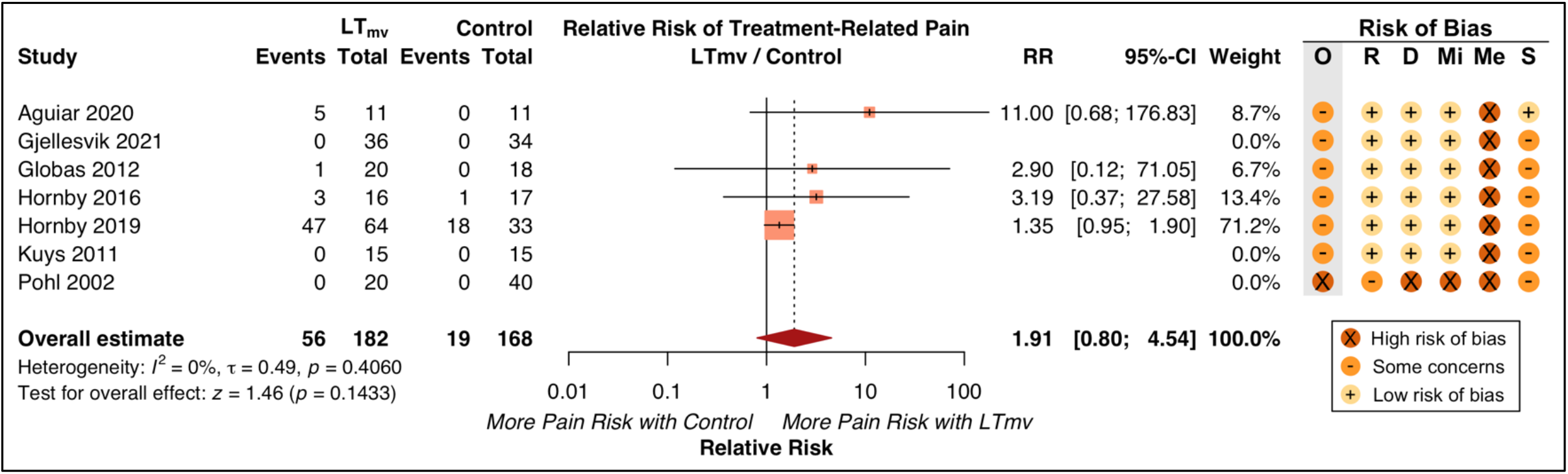
Relative risk (RR) of treatment-related pain for LT_mv_ versus control interventions. Results from a random effects meta-analysis. Only studies with some reporting about treatment-related pain are shown. Studies reporting no events in either group do not contribute any information about relative risk between groups. Risk-of-bias assessment was performed using the RoB-2 tool, yielding an overall (O) judgment based on the following domains: R, Bias arising from the randomization process; D, Bias due to deviations from intended intervention; Mi, Bias due to missing outcome data; Me, Bias in measurement of the outcome; S, Bias in selection of the reported result. LT_mv_, moderate-to-vigorous intensity locomotor training; Total, number of participants exposed to at least some of the intervention; Weight, % contribution of each study to the overall pooled estimate; I^2^, % of variance attributed to between-study heterogeneity; τ, estimated between-study standard deviation; Heterogeneity p-values are from Cochran’s Q test.

**Figure S7.**
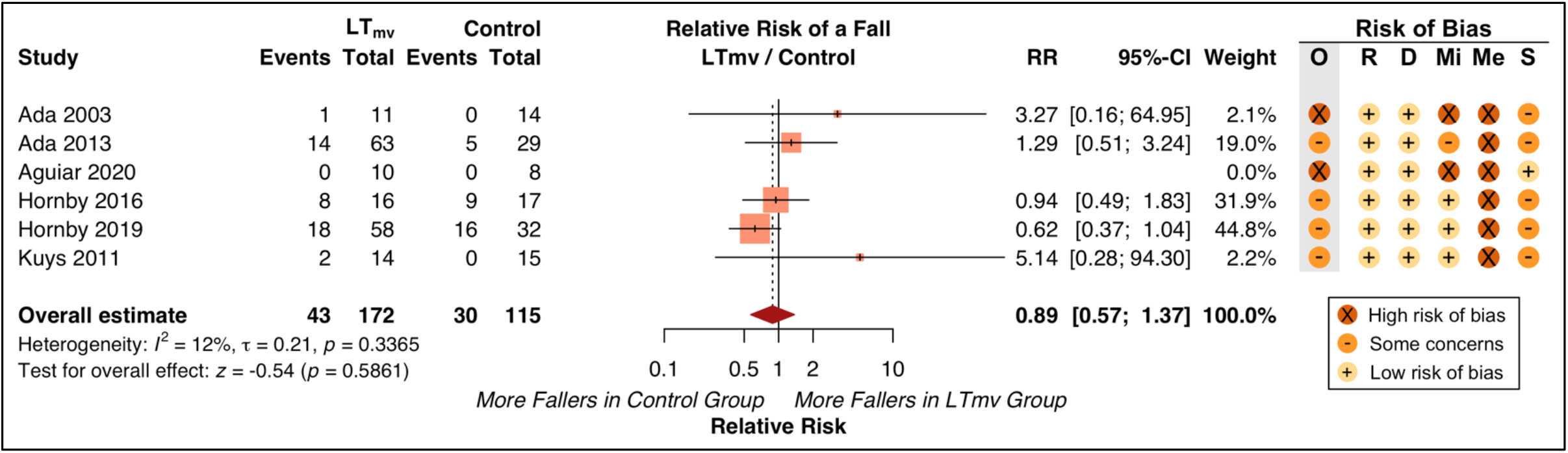
Relative risk (RR) of a fall for LT_mv_ versus control interventions. Results from a random effects meta-analysis. Only studies with some reporting about falls are shown. Studies reporting no fallers in either group do not contribute any information about relative risk between groups. Risk-of-bias assessment was performed using the RoB-2 tool, yielding an overall (O) judgment based on the following domains: R, Bias arising from the randomization process; D, Bias due to deviations from intended intervention; Mi, Bias due to missing outcome data; Me, Bias in measurement of the outcome; S, Bias in selection of the reported result. LT_mv_, moderate-to-vigorous intensity locomotor training; Total, number of participants with reported data; Weight, % contribution of each study to the overall pooled estimate; I^2^, % of variance attributed to between-study heterogeneity; τ, estimated between-study standard deviation; Heterogeneity p-values are from Cochran’s Q test.

**Figure S8.**
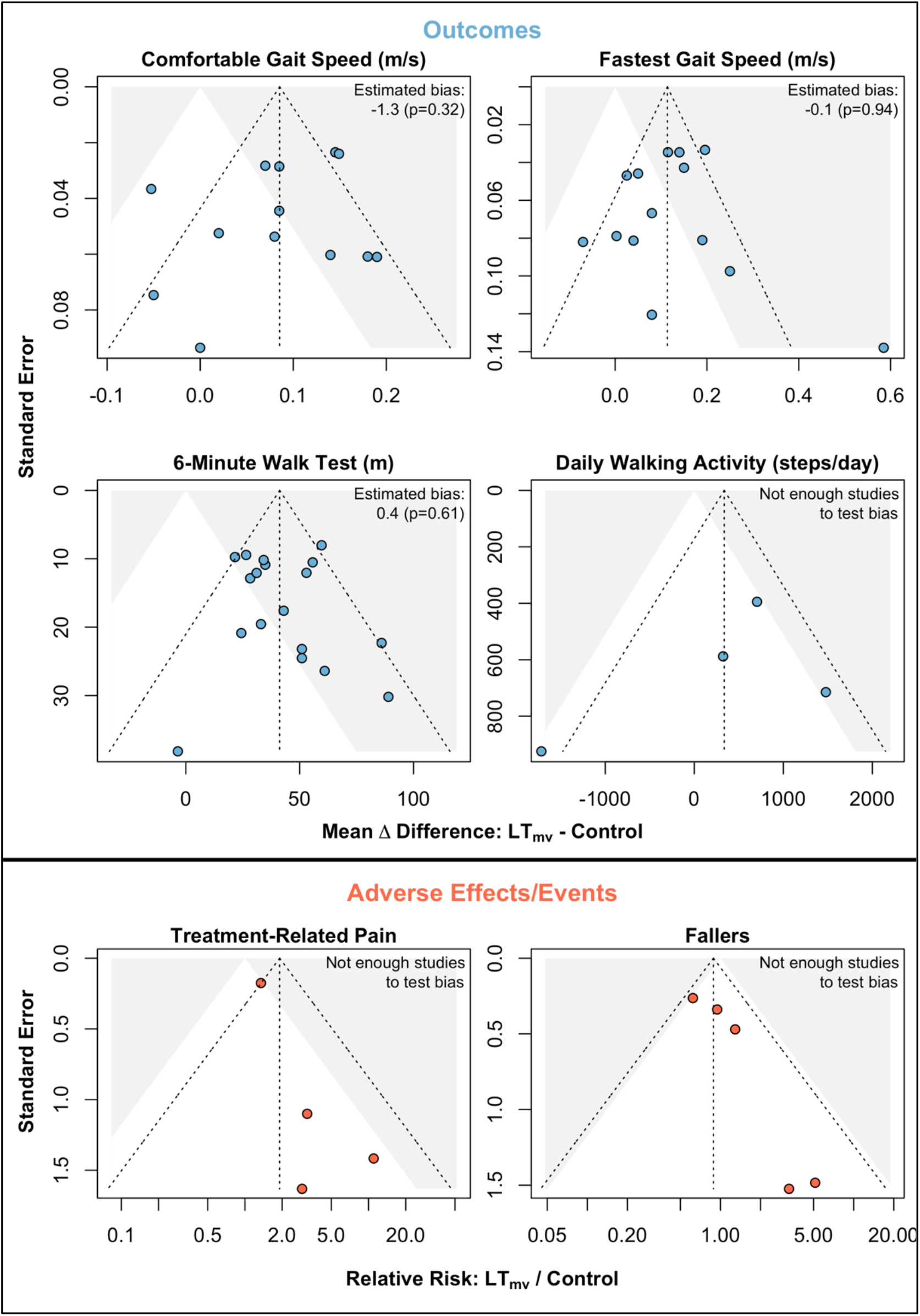
Publication bias assessment. Points are individual study estimates and vertical dotted lines show the pooled estimate from the random effects model. Shaded areas show the significance region (p<0.05) for the null hypothesis of no LT_mv_ versus control difference. The expected manifestation of publication bias is an asymmetrical distribution of studies around the pooled estimate, with smaller studies (higher standard error; lower on y-axis) being more likely to be reported if they have statistically significant findings. This would result in smaller studies being disproportionately clustered in the significance region to the right of the pooled estimate, with missing (non-reported) small studies in the non-significance region and to the left of the pooled estimate. Bias estimates are weighted regression coefficients testing the linear association between standard error and effect size, where significant positive values (not present here) would suggest publication bias.

**Figure S9.**
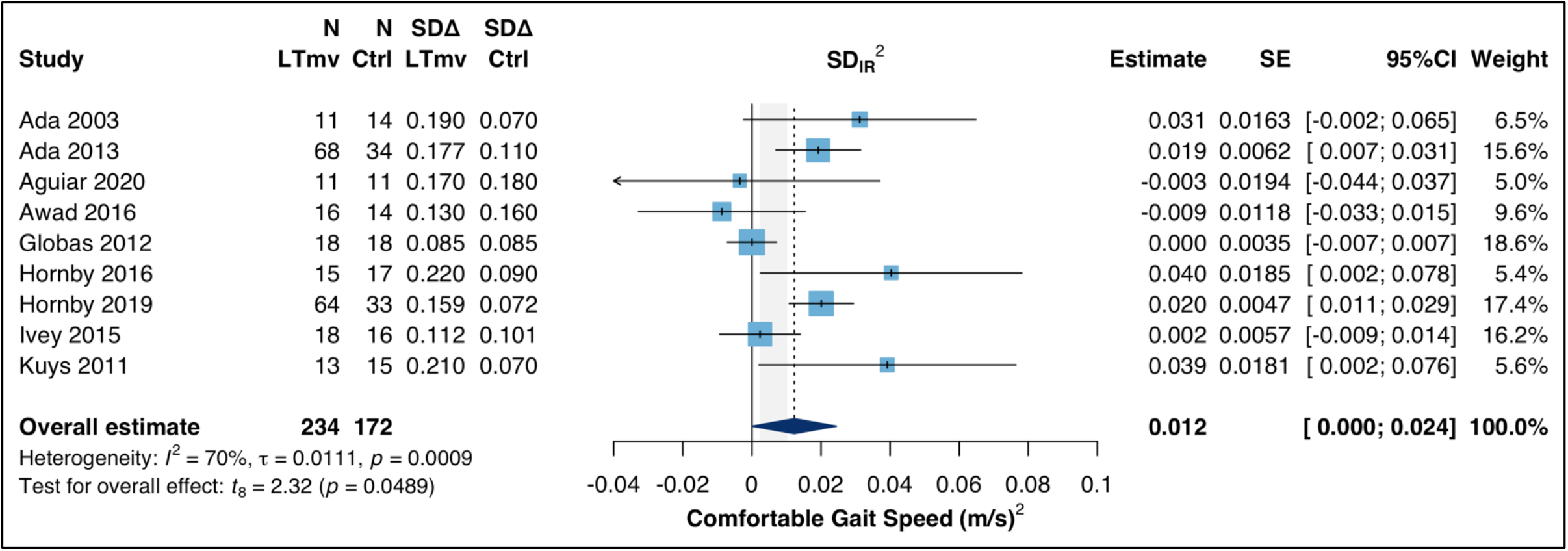
Variance of individual response (SD_IR2_): Comfortable gait speed. Results from a random effects meta-analysis. SD_IR_ estimates and 95%CI can be obtained by taking the square root of the values in this figure. The gray shaded region in the forest plot shows a range of clinically important difference thresholds in the literature (0.1-0.2 m/s), converted to SD_IR2_ units by halving and squaring (0.0025-0.01 [m/s]^2^). N, number of participants analyzed; SDΔ, standard deviation of change; LT_mv_, moderate-to-vigorous intensity locomotor training; SE, standard error; Weight, % contribution of each study to the overall pooled estimate; I^2^, % of variance attributed to between-study heterogeneity; τ, estimated between-study standard deviation; Heterogeneity p-values are from Cochran’s Q test.

**Figure S10.**
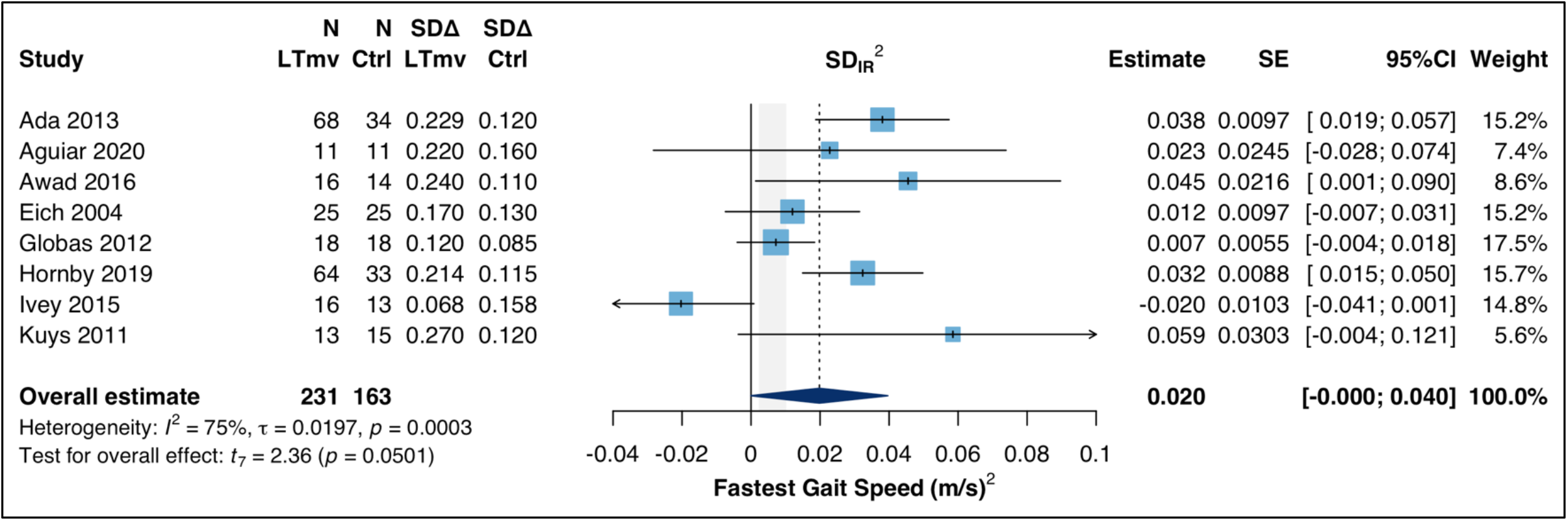
Variance of individual response (SD_IR2_): Fastest gait speed. Results from a random effects meta-analysis. SD_IR_ estimates and 95%CI can be obtained by taking the square root of the values in this figure. The gray shaded region in the forest plot shows a range of clinically important difference thresholds in the literature (0.1-0.2 m/s), converted to SD_IR2_ units by halving and squaring (0.0025-0.01 [m/s]^2^). N, number of participants analyzed; SDΔ, standard deviation of change; LT_mv_, moderate-to-vigorous intensity locomotor training; SE, standard error; Weight, % contribution of each study to the overall pooled estimate; I^2^, % of variance attributed to between-study heterogeneity; τ, estimated between-study standard deviation; Heterogeneity p-values are from Cochran’s Q test.

**Figure S11.**
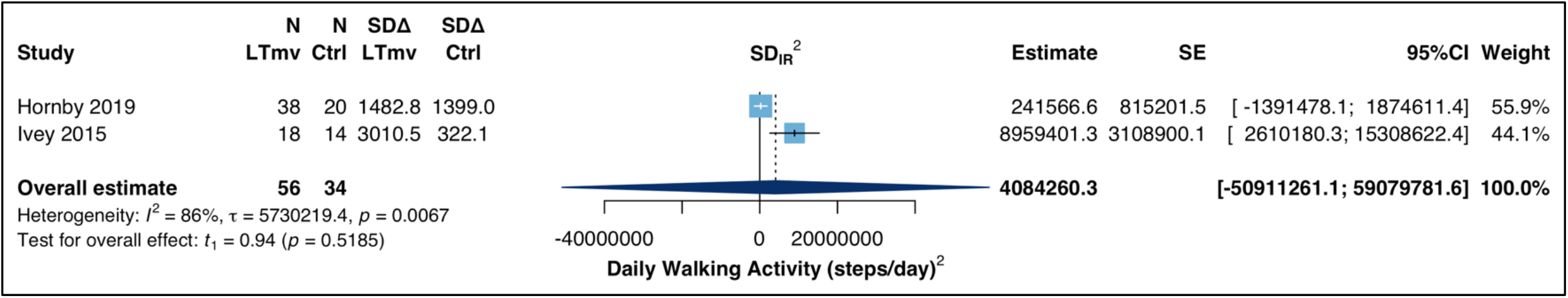
Variance of individual response (SD_IR2_): Daily walking activity. Results from a random effects meta-analysis. SD_IR_ estimates and 95%CI can be obtained by taking the square root of the values in this figure. N, number of participants analyzed; SDΔ, standard deviation of change; LT_mv_, moderate-to-vigorous intensity locomotor training; SE, standard error; Weight, % contribution of each study to the overall pooled estimate; I^2^, % of variance attributed to between-study heterogeneity; τ, estimated between-study standard deviation; Heterogeneity p-values are from Cochran’s Q test.

**Figure S12.**
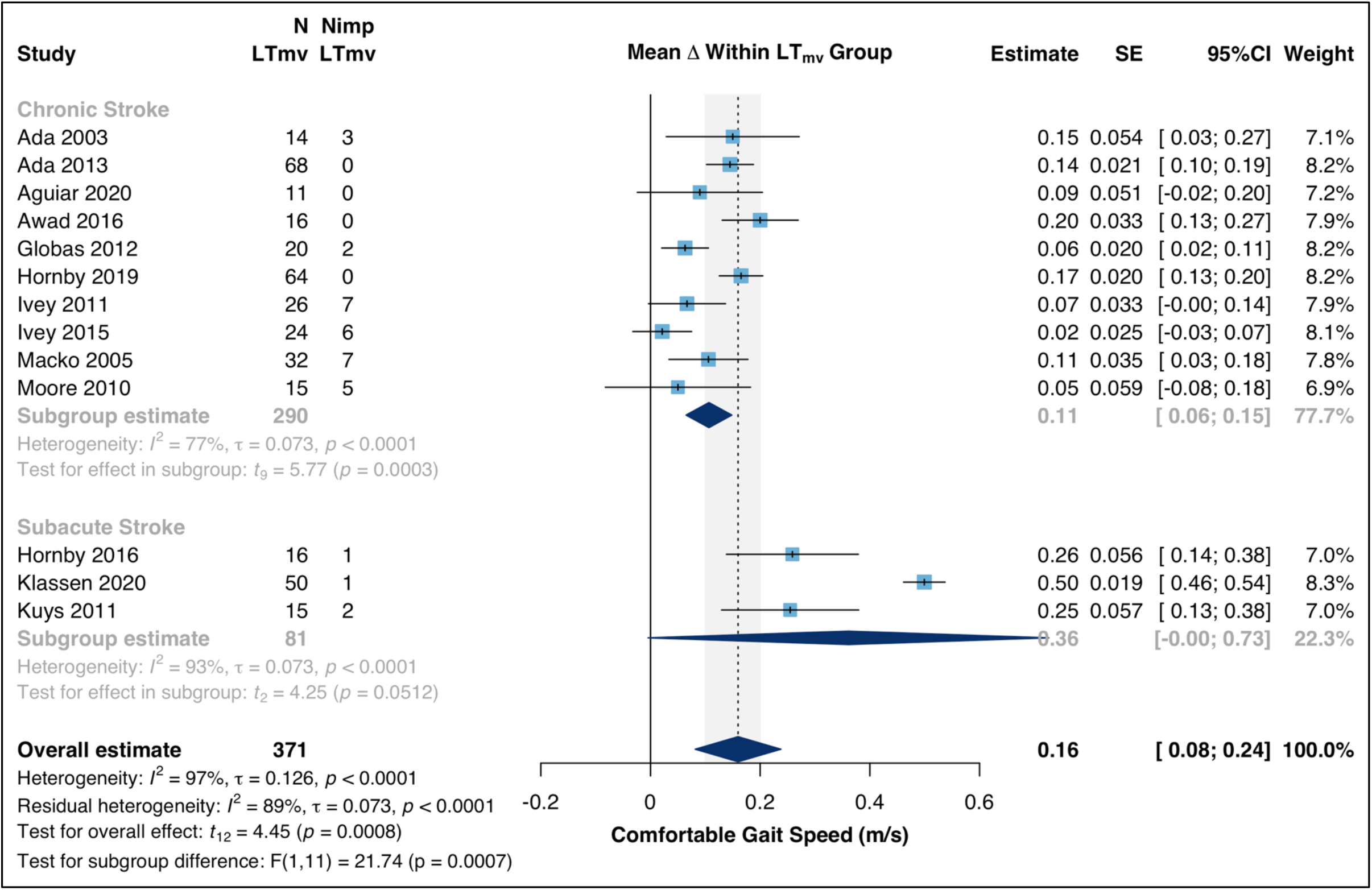
Mean changes (ι1) within LT_mv_ groups: Comfortable gait speed. Results from a random (mixed) effects meta-analysis stratified by stroke chronicity. The gray shaded region in the forest plot shows a range of clinically important difference thresholds in the literature (0.1-0.2 m/s). N, number of participants randomized; N_imp_, number of randomized participants not included in the reported analysis whose outcomes were imputed; LT_mv_, moderate-to-vigorous intensity locomotor training; SE, standard error; Weight, % contribution of each study and subgroup to the overall pooled estimate; I^2^, % of variance attributed to between-study heterogeneity; τ, estimated between-study standard deviation; Heterogeneity p-values are from Cochran’s Q test.

**Figure S13.**
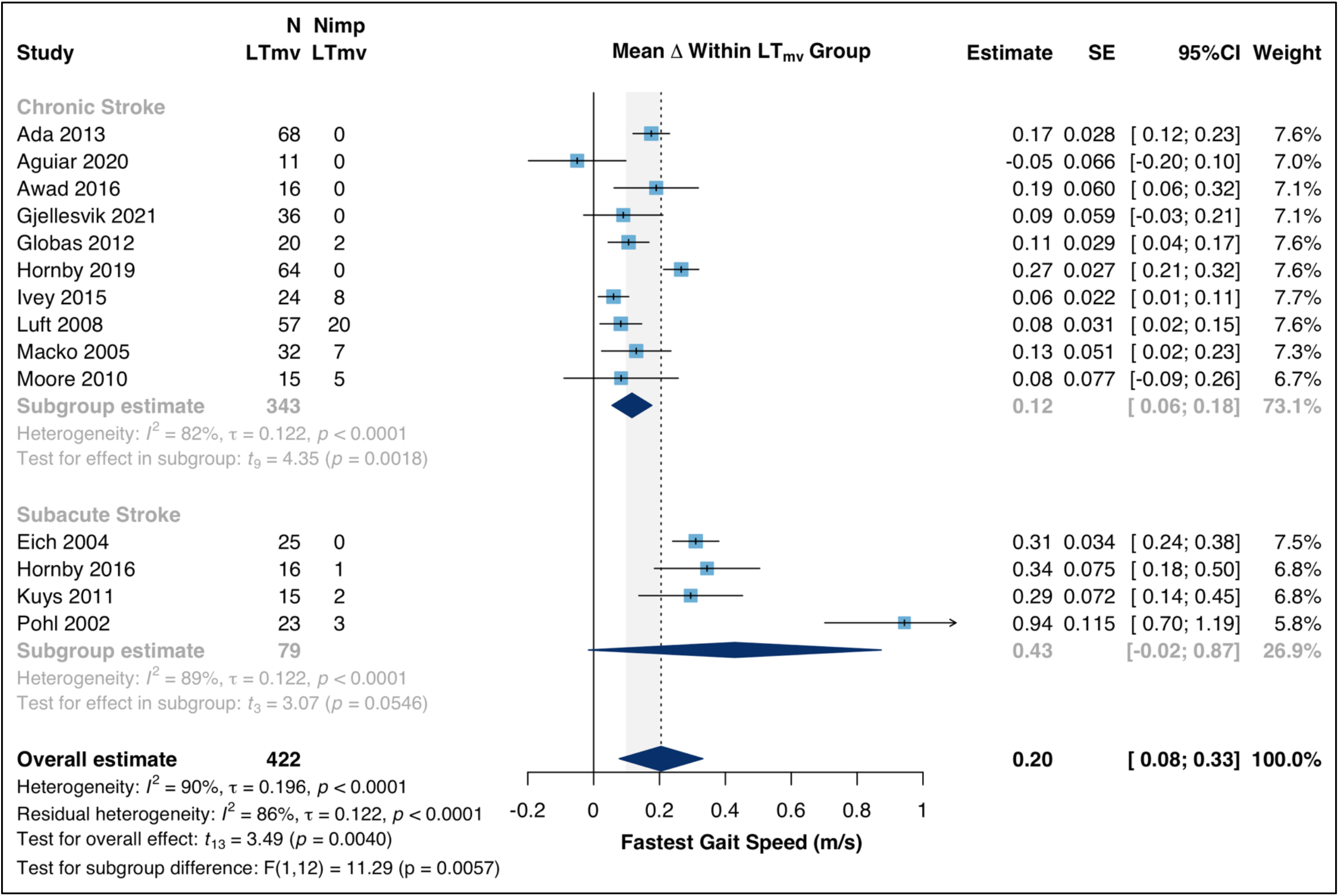
Mean changes (ι1) within LT_mv_ groups: Fastest gait speed. Results from a random (mixed) effects meta-analysis stratified by stroke chronicity. The gray shaded region in the forest plot shows a range of clinically important difference thresholds in the literature (0.1-0.2 m/s). N, number of participants randomized; N_imp_, number of randomized participants not included in the reported analysis whose outcomes were imputed; LT_mv_, moderate-to-vigorous intensity locomotor training; SE, standard error; Weight, % contribution of each study and subgroup to the overall pooled estimate; I^2^, % of variance attributed to between-study heterogeneity; τ, estimated between-study standard deviation; Heterogeneity p-values are from Cochran’s Q test.

**Figure S14.**
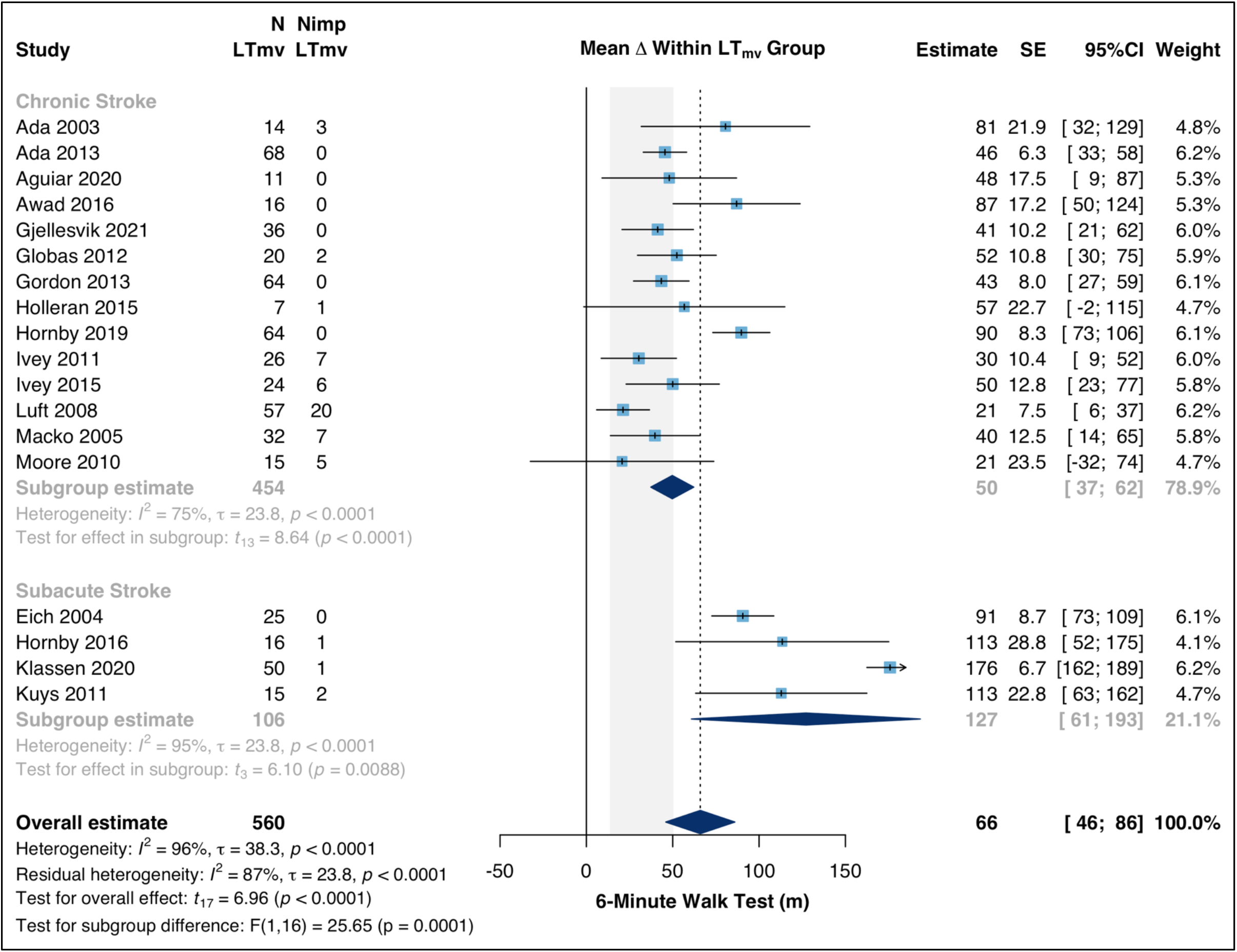
Mean changes (ι1) within LT_mv_ groups: 6-minute walk test. Results from a random (mixed) effects meta-analysis stratified by stroke chronicity. The gray shaded region in the forest plot shows a range of clinically important difference thresholds in the literature (14-50 m). N, number of participants randomized; N_imp_, number of randomized participants not included in the reported analysis whose outcomes were imputed; LT_mv_, moderate-to-vigorous intensity locomotor training; SE, standard error; Weight, % contribution of each study and subgroup to the overall pooled estimate; I^2^, % of variance attributed to between-study heterogeneity; τ, estimated between-study standard deviation; Heterogeneity p-values are from Cochran’s Q test.

**Figure S15.**
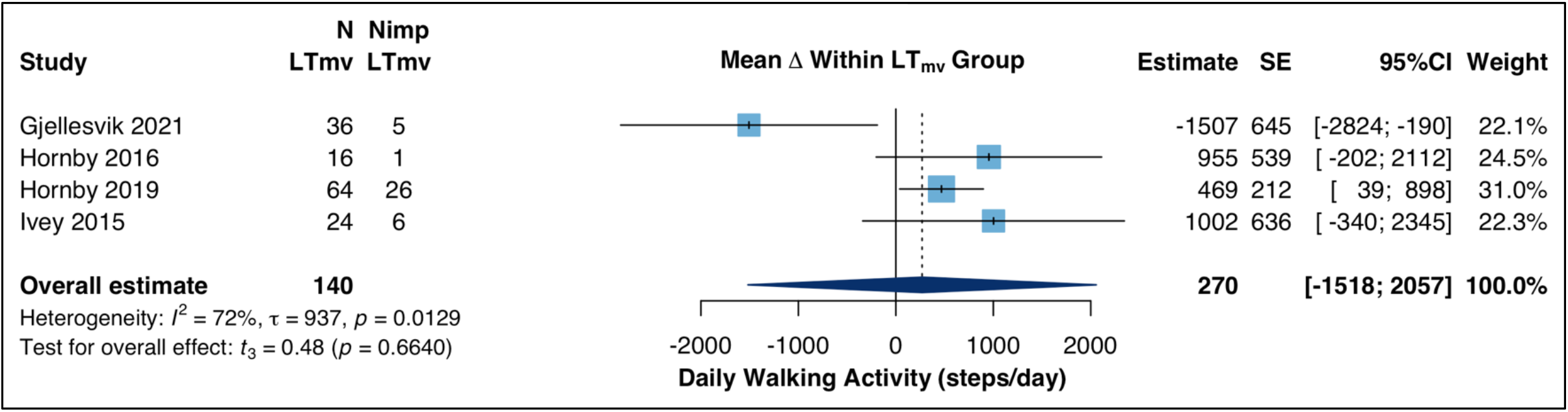
Mean changes (ι1) within LT_mv_ groups: Daily walking activity. Results from a random effects meta-analysis. N, number of participants randomized; N_imp_, number of randomized participants not included in the reported analysis whose outcomes were imputed; LT_mv_, moderate-to-vigorous intensity locomotor training; SE, standard error; Weight, % contribution of each study to the overall pooled estimate; I^2^, % of variance attributed to between-study heterogeneity; τ, estimated between-study standard deviation; Heterogeneity p-values are from Cochran’s Q test.

**Figure S16.**
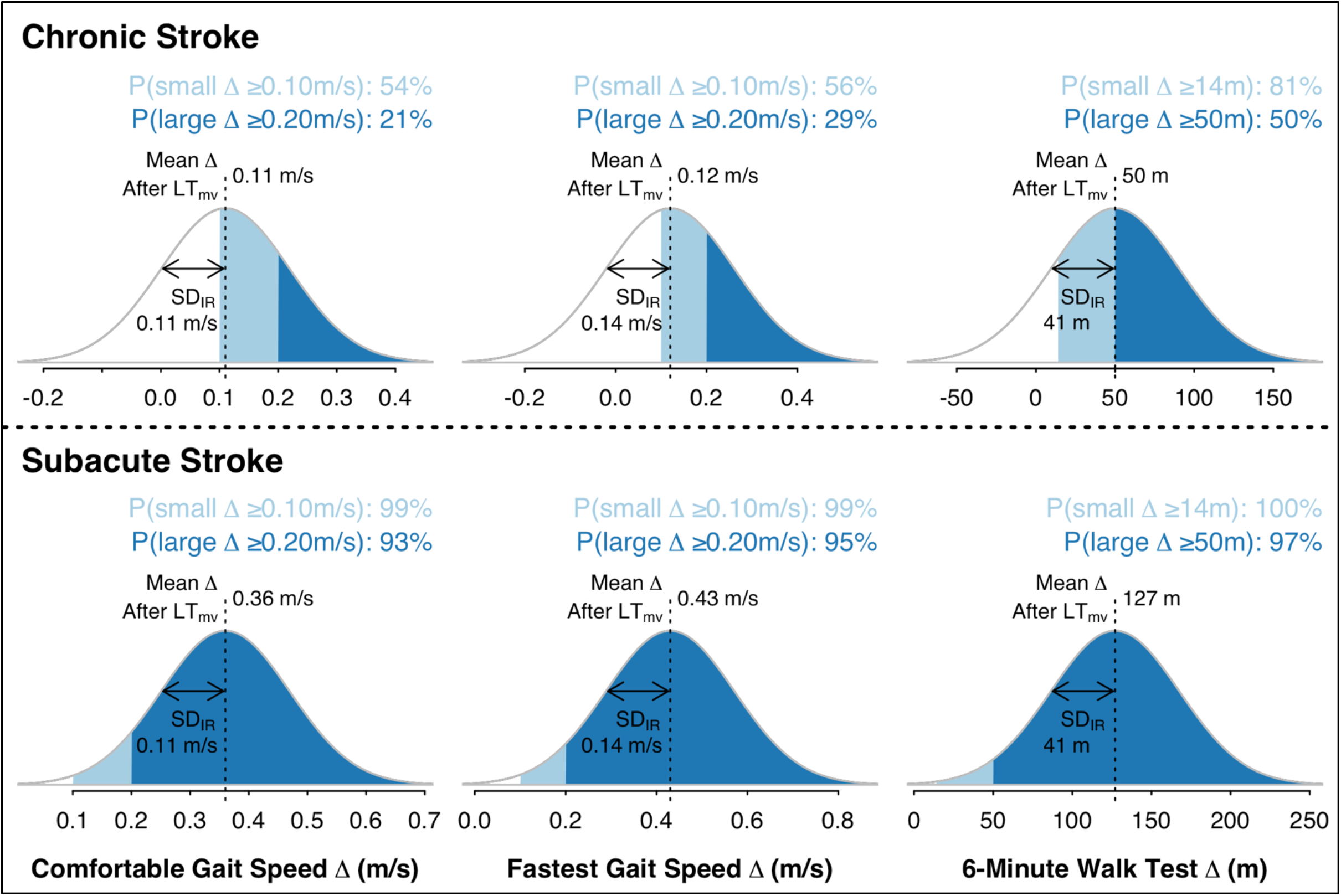
Estimated proportions of participants with meaningful gains during LT_mv_. Graphs show the estimated distributions of individual changes (Δ) during LT_mv_, based on the meta-analysis results. Separate distributions were calculated for each outcome measure (from left to right: comfortable gait speed, fastest gait speed, 6-minute walk test) and for each stroke chronicity subgroup (top panel, chronic; bottom panel, subacute). The mean of each distribution is the meta-analysis estimate of the mean within LT_mv_ groups, and the SD of each distribution is the meta-analysis estimate of the SD_IR_ (the estimate of the true individual variability in responsiveness). Reported probabilities (P) are the % of individual scores that were at or above different clinically important difference thresholds from the literature. These thresholds ranged from 0.10 m/s (small) to 0.20 m/s (large) for gait speed^28-31^ and from 14 m (small) to 50 m (large) for the 6-minute walk test.

## APPENDIX Example Search Strategy (PubMed)

1. Stroke [MeSH Terms] OR cerebrovascular accident OR cva OR hemipleg* OR hemipar* OR brain infarction OR brain stem infarction OR cerebral infarction OR subcortical infarction OR cerebral hemorrhage OR brain hemorrhage OR brain stem hemorrhage OR subcortical hemorrhage OR interventricular hemorrhage OR subarachnoid hemorrhage OR intracerebral hemorrhage
2. Walk* OR gait OR ambulat* OR locomot* OR treadmill OR overground OR step*
3. Exercis* OR train* OR program OR rehabilitat* OR intervention
4. NOT (animals[MeSH Terms] NOT humans[MeSH Terms])
5. NOT “meta-analysis”[Publication Type] NOT “systematic review”[Publication Type] NOT “review”[Publication Type]
6. ”randomized controlled trial”[Publication Type] AND “clinical trial”[Publication Type])
7. 1995/01/01:2022/03/01[Date -Publication]

#1 AND #2 AND #3 AND #4 AND #5 AND #6 AND #7

